# Exploring real-world user experiences of GLP-1 receptor agonist therapy for obesity treatment, and barriers and motivators to adherence

**DOI:** 10.64898/2025.12.11.25342052

**Authors:** Alex Griffiths, Oliver M Shannon, Marie Spreckley, Kate Austin, Ellen Fallows, Ken Clare, Louisa Ells, Jamie Matu

## Abstract

**Background:** Glucagon-like peptide-1 receptor agonists (GLP-1RAs) demonstrate efficacy for weight loss in clinical trials, yet real-world implementation challenges remain poorly understood. This study evaluated current and former user experiences of GLP-1RA medications for obesity treatment in the UK.

**Methods:** A cross-sectional online survey was administered via Prolific to 352 current and 272 former GLP-1RA users. Questionnaires were designed to gather data on 1) Participant characteristics, 2) Medication use, access and provision, 3) Motivations, experiences and perceived outcomes, 4) Healthcare provider support, 5) Barriers and motivators to adherence, and 6) Discontinuation and post-treatment impacts. Data were presented descriptively, as well as inferentially using chi-squared tests to compare differences in experiences between current and former users and between demographic subgroups.

**Results:** Current users were more likely than former users to report that the medication helped them achieve their goals (84% vs. 67%, p<0.001). Treatment was predominantly accessed privately online, but to a greater extent in current users (current: 76%, former: 58%, *p*<0.001). Healthcare support was generally reported as adequate overall (current: 62%, former: 54%, *p*=0.02) but rated consistently lower for dietary (current: 49%, former: 49%, *p*=0.79), physical activity (current: 36%, former: 37%, *p*=0.71), and psychological care (current: 27%, former: 28%, *p*=0.71). Cost emerged as the primary barrier to adherence and main reason for discontinuation (31%), disproportionately affecting lower socioeconomic groups. The prevalence of significant side effects was higher in former than current users (38% vs. 29%, *p*=0.02) and contributed to 25% of discontinuations. Post-discontinuation, 45% reported weight regain, 40% maintained weight, and 15% continued losing weight.

**Conclusion:** While GLP-1RA treatment effectively supported weight goals, sustainability is undermined by high costs, inadequate holistic support, and side-effect burden. Findings emphasize the need for integrated multidisciplinary care models with tailored approaches addressing distinct demographic barriers.

## Introduction

Obesity is a complex, chronic disease associated with several comorbidities, including type 2 diabetes [1] and cardiovascular disease [2]. Traditional approaches to managing obesity often focus on diet and physical activity changes [3], however adherence and long-term weight loss are typically poor [4]. Glucagon-like peptide-1 receptor agonists and combination incretin medications (hereafter referred to as GLP-1RAs) provide an effective pharmacological option for individuals who have struggled to achieve or sustain weight loss through behaviour-change interventions alone [5,6]. Whilst clinical trials consistently demonstrate the efficacy of GLP-1RAs for weight reduction [7], these trial populations do not reflect the diversity of the UK population, and may not capture the complexities of treatment access, clinical support, and post-discontinuation outcomes in routine practice. The American College of Lifestyle Medicine, the American Society for Nutrition, the Obesity Medicine Association, and The Obesity Society have recently issued joint advisory priorities to optimise GLP1-RA therapy. They emphasise the necessity of concomitant lifestyle and behavioural support to improve nutritional status, preserve lean mass, and promote long-term health [8]. However, there is potentially a disconnect between emerging clinical guidance and our understanding of the patient experience in relation to GLP1-RAs.

Recent UK research has begun to address these gaps by exploring community perspectives on GLP-1RAs, demonstrating high awareness but persistent concerns about safety, cost, and stigma that influence treatment initiation [9]. In addition, Naveed et al. [10] provided early insights into real-world GLP-1RA discontinuation in the UK, identifying cost and side effects as key reasons for stopping treatment. However, these studies focused on perceived factors influencing treatment initiation or reasons for discontinuation, rather than a comprehensive overview of the patient journey. Critical gaps in the evidence remain regarding real-world implementation. Specifically, there is a lack of understanding of (1) the comparative experiences of those who continue treatment (current users) versus those who discontinue (former users); (2) the prevalence and quality of healthcare support alongside treatment; (3) medication access and clinical oversight, which dictate the safety and feasibility of treatment in routine care; (4) post-discontinuation weight trajectories and behavioural outcomes to inform follow-up care and (5) demographic differences across all domains, given GLP-1RA treatment requires personalised rather than uniform approaches (Reilly-Harrington et al., 2025).

Therefore, this study aimed to evaluate current and former user experiences of GLP-1RA medications for obesity treatment, with particular emphasis on identifying barriers and motivators to use, and reasons for discontinuation and post-treatment responses. These data may inform strategies to optimise the long-term effectiveness of GLP-1RAs in obesity management and guide the development of more tailored, person-centred care pathways.

## Methods

### Study design

This study employed a cross-sectional, quantitative survey, administered via the online research platform Prolific.

### Participants

Inclusion criteria required participants to be UK adults (aged 18 or older) with current or former experience using GLP1-RAs for obesity treatment. To focus specifically on obesity related use, those who were prescribed GLP1-RAs primarily for diabetes management were ineligible for inclusion in this study. Participants were provided modest remuneration for their time (∼£12/h pro-rata).

### Questionnaire development and administration

Two separate questionnaires were designed to assess the experiences of 1) current users of GLP-1RA medication and 2) former users who had discontinued treatment. Both questionnaires aimed to quantify treatment experiences, barriers and motivators to adherence, and reasons and responses to discontinuation. The content of the questionnaires were informed by the COM-B model [11] and the Theoretical Domains Framework [12]. The questionnaires were developed, pilot tested, and subsequently edited working with individuals with a lived experience of obesity and GLP1-RA use to ensure the inclusion of appropriate content and language. The questionnaires were structured into the following sections: 1) Demographics, 2) User experiences of GLP1-RAs for obesity treatment, and 3) Factors influencing adherence to GLP1-RAs and behaviour change support. Following review of the data, our findings were logically presented to improve the clarity of our results section as follows: 1) Participant characteristics, 2) Medication use, access and provision, 3) Motivations, experiences and perceived outcomes, 4) Healthcare provider support, 5) Barriers and motivators to adherence, and 6) Discontinuation and post-treatment impacts. Healthcare provider support refers to the support provided by the medication prescriber.

### Statistical analysis

Data were analysed using SPSS version 29. Data were reported descriptively for both cohorts, as well as a comparison between the two (i.e. current vs. former users). Reasons for discontinuation and post-treatment impacts were evaluated and reported descriptively in former users only. Differences in responses between the two cohorts as well as differences in population sub-groups were explored using a chi squared test where appropriate. These population subgroups were defined as age (younger (<40 years) vs. older (≥40 years)), gender (males vs. females), current BMI (lower (≤30 kg/m^2^) vs. higher (>30 kg/m^2^)), BMI at the start of treatment (lower (25-39.9 kg/m^2^) vs. higher (≥40 kg/m^2^), ethnicity (white vs other ethnic groups), education (higher (undergraduate degree, Master’s degree or PhD) vs. lower (GCSE, A Level, vocational, other)), self-defined social status [13] (lower (1-5 ladder scale) vs higher (6-10 ladder scale) and exercise participation (yes vs. no).

## Results

### Participant characteristics

Participants included 352 current and 272 former GLP-1RA users. Participant characteristics were broadly similar between current and former users (Table 1). Most participants were white, female, and of moderate self-defined social status, with the majority in the 30–39-year age group and holding at least an undergraduate degree. A higher proportion of current users were living with obesity both at survey and treatment initiation compared with former users.

**Table 1.** Participant characteristics of current and former GLP-1 agonist users.

| Participant Characteristics | Current Users (n = 352) |  | Former users (n = 272) |  |
| --- | --- | --- | --- | --- |
|  | n | % | n | % |
| **Age** |  |  |  |  |
| 18–29 | 53 | 15 | 56 | 21 |
| 30–39 | 119 | 34 | 93 | 34 |
| 40–49 | 82 | 23 | 58 | 21 |
| 50–59 | 67 | 19 | 42 | 15 |
| 60–69 | 30 | 9 | 22 | 8 |
| 70 or over | 1 | <1 | 1 | <1 |
| **Gender** |  |  |  |  |
| Female | 268 | 76 | 186 | 68 |
| Male | 83 | 24 | 84 | 31 |
| Non-binary/Other | 1 | <1 | 2 | 1 |
| **BMI at time of survey** |  |  |  |  |
| <18.5 | 0 | 0 | 1 | <1 |
| 18.5–24.9 | 45 | 13 | 60 | 22 |
| 25–29.9 | 116 | 33 | 110 | 40 |
| 30–39.9 | 147 | 42 | 76 | 28 |
| 40 or over | 44 | 13 | 25 | 9 |
| **BMI at start of treatment** |  |  |  |  |
| 25–29.9 | 32 | 9 | 53 | 19 |
| 30–39.9 | 196 | 56 | 154 | 57 |
| 40 or over | 124 | 35 | 65 | 24 |
| **Ethnicity** |  |  |  |  |
| White | 311 | 88 | 230 | 85 |
| Asian or Asian British | 12 | 3 | 10 | 4 |
| Black | 19 | 5 | 23 | 8 |
| Mixed | 10 | 3 | 7 | 3 |
| Other | 0 | 0 | 2 | 1 |
| **Region** |  |  |  |  |
| North East | 25 | 7 | 21 | 8 |
| North West | 39 | 11 | 36 | 13 |
| Yorkshire | 25 | 7 | 28 | 10 |
| East Midlands | 35 | 10 | 17 | 6 |
| West Midlands | 24 | 7 | 19 | 7 |
| South East | 77 | 22 | 33 | 12 |
| South West | 23 | 7 | 22 | 8 |
| London | 34 | 10 | 43 | 16 |
| Scotland | 39 | 11 | 30 | 11 |
| Wales | 24 | 7 | 13 | 5 |
| Northern Ireland | 7 | 2 | 10 | 4 |
| **Highest level of education** |  |  |  |  |
| O Levels/GCSE | 33 | 9 | 30 | 11 |
| A Level or equivalent | 39 | 11 | 28 | 10 |
| Vocational qualification | 51 | 14 | 36 | 13 |
| Undergraduate degree | 159 | 45 | 117 | 43 |
| Master's degree | 62 | 18 | 50 | 18 |
| Doctorate | 6 | 2 | 8 | 3 |
| Other | 2 | 1 | 3 | 1 |
| **Self-defined social status** |  |  |  |  |
| Lower (1-3) | 27 | 8 | 23 | 8 |
| Moderate (4-7) | 287 | 81 | 212 | 78 |
| Higher (8-10) | 38 | 11 | 37 | 14 |
| **Participates in PA** |  |  |  |  |
| Yes | 242 | 69 | 196 | 72 |
| No | 110 | 31 | 76 | 28 |
| **Moderate-intensity PA** |  |  |  |  |
| <60 minutes per week | 52 | 21 | 30 | 15 |
| 60–90 minutes per week | 76 | 31 | 56 | 29 |
| 90–120 minutes per week | 61 | 25 | 49 | 25 |
| 120–150 minutes per week | 23 | 10 | 19 | 10 |
| >150 minutes per week | 30 | 12 | 42 | 21 |
| **Vigorous-intensity PA** |  |  |  |  |
| <60 minutes per week | 170 | 70 | 113 | 58 |
| 60–90 minutes per week | 44 | 18 | 42 | 21 |
| 90–120 minutes per week | 14 | 6 | 14 | 7 |
| 120–150 minutes per week | 6 | 2 | 15 | 8 |
| >150 minutes per week | 8 | 3 | 12 | 6 |
| **Long-term health conditions** |  |  |  |  |
| Heart or circulatory | 20 | 6 | 18 | 7 |
| Metabolic condition | 50 | 14 | 35 | 13 |
| Neurodegenerative disease | 5 | 1 | 0 | 0 |
| Kidney disease | 1 | <1 | 3 | 1 |
| Gastrointestinal disorders | 12 | 3 | 18 | 7 |
| Lung disease | 7 | 2 | 5 | 2 |
| Anxiety or depression | 158 | 45 | 120 | 44 |
| Other | 51 | 14 | 25 | 9 |
| None | 132 | 38 | 107 | 39 |

BMI – Body Mass Index, PA – Physical Activity

### Medication use, access and provision

Full reporting of data related to medication, access, and provision are included in Supplementary Material 1. Current users predominantly used Tirzepatide (80% vs 46% of former users), while former users mostly used Semaglutide (49% vs 19% of current users; p<0.001). Most current users obtained prescriptions privately, typically online (76%), compared with 58% of former users (*p*<0.001). NHS provision was less common in current and former groups, via both online (4% vs. 7%) or face-to-face (16% vs. 23%) routes (*p*<0.001). Current users were also more likely to self-weigh to confirm eligibility (76% vs. 61%, *p*<0.001), with a greater proportion providing proof via photo or video (76% vs. 64%, p=0.01). Finally, more current users reported that their GP was aware of their treatment (70% vs. 56%, p<0.001).

### Motivations, experiences and perceived outcomes of using GLP-1 agonist medication

Current users were more likely than former users to report that the medication helped them achieve their goals (84% vs. 67%, p<0.001), though both groups noted that effectiveness generally declined over time (49% vs. 45%, p=0.51). Most participants reported that GLP-1RA medication was easy to use (96% vs. 93%) and access (71% vs. 74%). Around one third reported significant side effects, with fewer current users affected (29% vs. 38%, p=0.02). Current users were also more likely to report better perceived outcomes than former users, with improvements in quality of life (87% vs. 58%), mental health (58% vs. 40%), and perceived consumption of less (96% vs. 92%), yet healthier (78% vs. 68%) food (all p≤0.046).

Participants in the current user group also reported increased prevalence of benefits experienced including weight loss (94% vs. 83%), body image (72% vs. 53%), self-confidence (68% vs. 52%), improved health outcomes (53% vs. 30%), quality of life (52% vs. 33%), diet quality (56% vs. 40%), physical activity (45% vs. 28%), pain management (23% vs. 7%), social life (17% vs. 14%) and reduction in other medications (8% vs. 3%). Supplement use was common in current and former groups, most frequently multivitamins (66% vs. 54%), vitamin D (37% vs. 35%) and protein powders (19% vs. 19%). Most participants reported decreased enjoyment of food while using the medication (62% vs. 68%, p=0.18). A broader summary of experiences and perceived benefits is provided in Supplementary Material 2.

### Healthcare provider support

Participant experiences of healthcare provider support varied between groups (Supplementary Material 3). A higher percentage of current users reported adequate overall support compared with former users (62% vs. 54%, p=0.02), and more current users agreed that their healthcare provider was knowledgeable about GLP-1RA medication (74% vs. 67%, p<0.001) and regularly monitored weight (61% vs. 52%, p=0.048). Around half of participants in both groups felt they received adequate dietary advice (49% vs. 49%, p=0.79), while fewer current and former users reported sufficient support for physical activity (36% vs. 37%, p=0.71) or psychological needs (27% vs. 28%, p=0.71). Most participants felt comfortable discussing concerns with their healthcare provider (72% vs. 67%, p=0.28) and generally adhered to the advice provided (94% vs. 96%, p=0.39). Healthcare support was reported to make little difference to adherence for most participants (1% vs. 5%, p=0.02).

### Barriers and motivators for adherence

The most frequently reported barrier to adherence (Table 2) was cost of medication, with around a third of current and former users reporting it as extremely challenging. Side effects were a greater barrier for former users, while current users more often reported social stigma related to GLP1-RA use as a challenge. In contract, access to medication and social life impacts were generally not perceived as barriers.

**Table 2.**
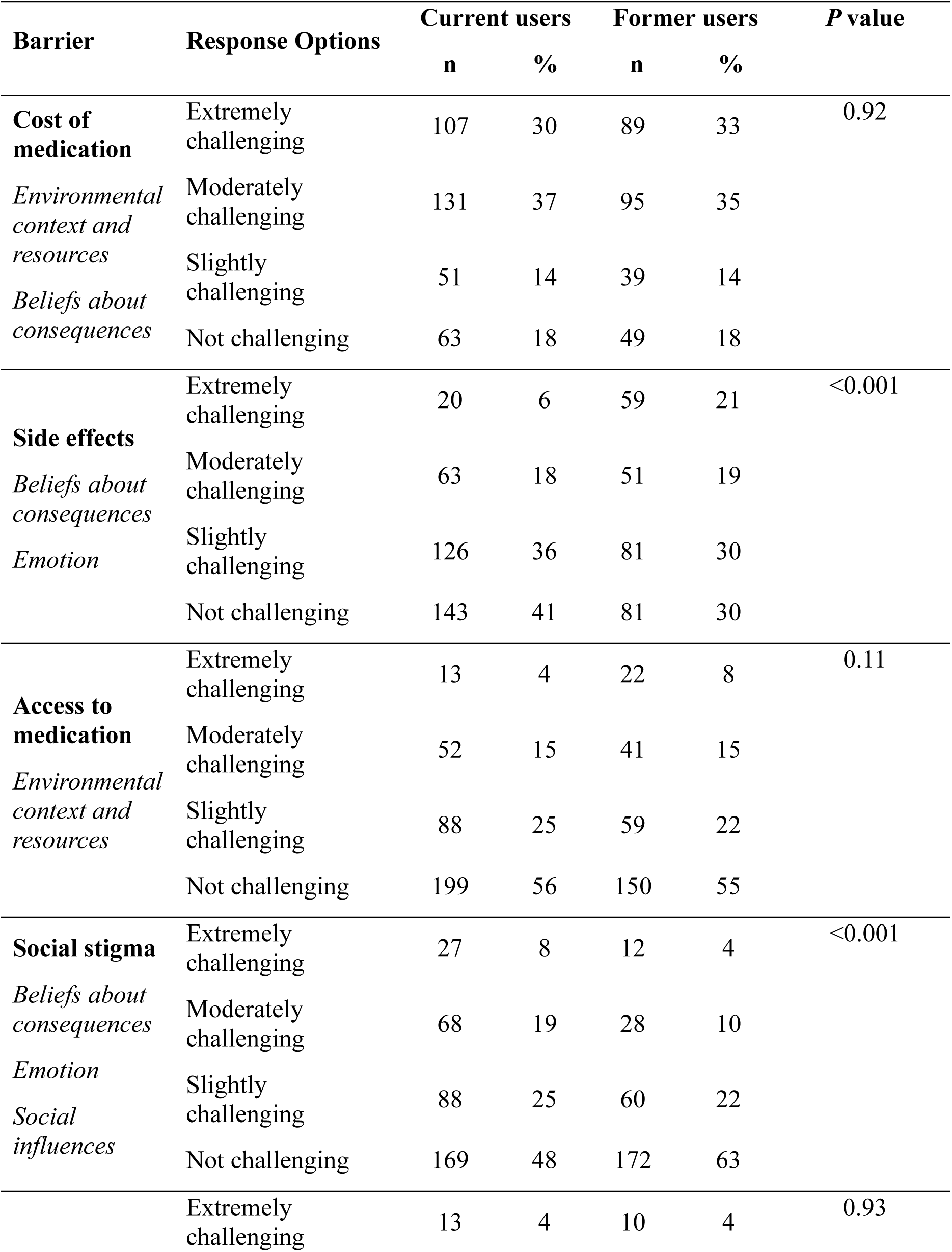
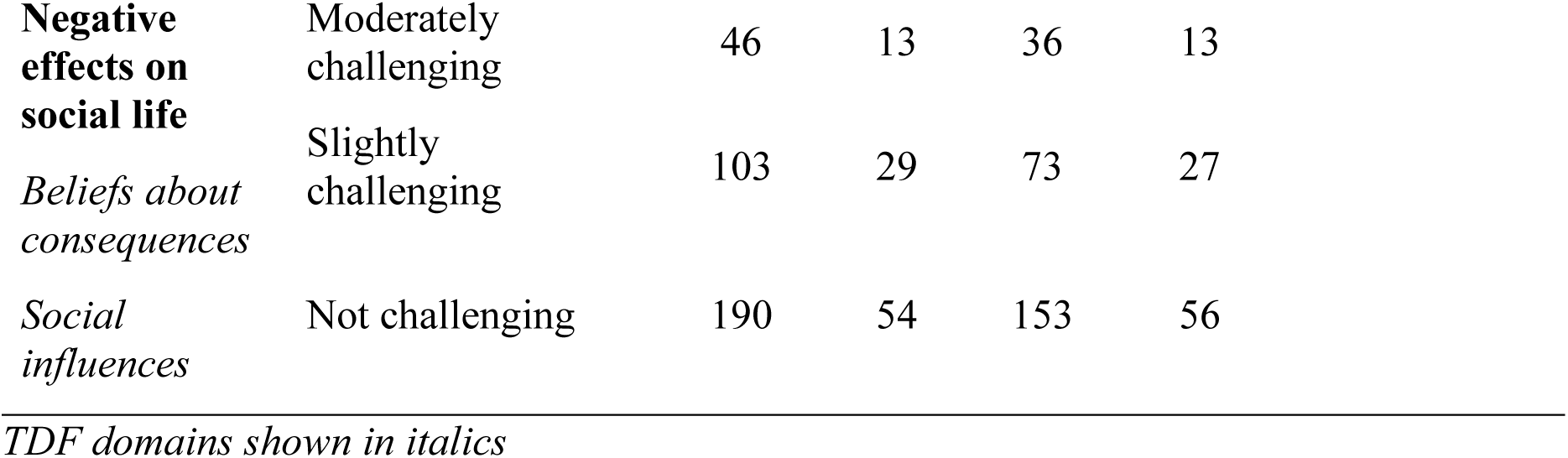
Barriers to GLP-1 receptor agonist medication adherence.

| Barrier | Response Options | Current users |  | Former users |  | *P* value |
| --- | --- | --- | --- | --- | --- | --- |
|  |  | n | % | n | % |  |
| **Cost of medication** | Extremely challenging | 107 | 30 | 89 | 33 | 0.92 |
| *Environmental context and resources* | Moderately challenging | 131 | 37 | 95 | 35 |  |
| *Beliefs about consequences* | Slightly challenging | 51 | 14 | 39 | 14 |  |
|  | Not challenging | 63 | 18 | 49 | 18 |  |
| **Side effects** | Extremely challenging | 20 | 6 | 59 | 21 | <0.001 |
| *Beliefs about consequences* | Moderately challenging | 63 | 18 | 51 | 19 |  |
| *Emotion* | Slightly challenging | 126 | 36 | 81 | 30 |  |
|  | Not challenging | 143 | 41 | 81 | 30 |  |
| **Access to medication** | Extremely challenging | 13 | 4 | 22 | 8 | 0.11 |
| *Environmental context and resources* | Moderately challenging | 52 | 15 | 41 | 15 |  |
|  | Slightly challenging | 88 | 25 | 59 | 22 |  |
|  | Not challenging | 199 | 56 | 150 | 55 |  |
| **Social stigma** | Extremely challenging | 27 | 8 | 12 | 4 | <0.001 |
| *Beliefs about consequences* | Moderately challenging | 68 | 19 | 28 | 10 |  |
| *Emotion* | Slightly challenging | 88 | 25 | 60 | 22 |  |
| *Social influences* | Not challenging | 169 | 48 | 172 | 63 |  |
|  | Extremely challenging | 13 | 4 | 10 | 4 | 0.93 |

|  |  |  |  |  |  |
| --- | --- | --- | --- | --- | --- |
| **Negative effects on social life** | Moderately challenging | 46 | 13 | 36 | 13 |
| *Beliefs about consequences* | Slightly challenging | 103 | 29 | 73 | 27 |
| *Social influences* | Not challenging | 190 | 54 | 153 | 56 |

*TDF domains shown in italics*

Motivators to GLP-1 medication adherence generally differed, with current users identifying multiple factors as having greater influences on adherence (Table 3). The predominant motivators in both groups included weight loss, improved health outcomes, improved body image, increased self-confidence, and improved quality of life.

**Table 3.** Motivators to GLP-1 receptor agonist medication adherence.

| **Motivators** | **Response Options** | **Current users** |  | **Former users** |  |  |
| --- | --- | --- | --- | --- | --- | --- |
|  |  | **n** | **%** | **n** | **%** |  |
| **Improved health outcomes** | Extremely important | 237 | 67 | 136 | 50 | **<0.001** |
| *Beliefs about consequences* | Moderately important | 101 | 29 | 96 | 35 |  |
| *Goals* | Slightly important | 12 | 3 | 30 | 11 |  |
| *Emotion* | Not important | 2 | 1 | 10 | 4 |  |
| **Weight loss** | Extremely important | 306 | 87 | 217 | 80 | **0.03** |
| *Beliefs about consequences* | Moderately important | 43 | 12 | 46 | 17 |  |
| *Goals* | Slightly important | 3 | 1 | 7 | 3 |  |
| *Emotion* | Not important | 0 | 0 | 2 | 1 |  |
| **Improved body image** | Extremely important | 217 | 61 | 144 | 53 | **0.03** |
| *Beliefs about consequences* | Moderately important | 98 | 28 | 88 | 32 |  |
| *Goals* | Slightly important | 31 | 9 | 26 | 10 |  |
| *Emotion* |  |  |  |  |  |  |
| *Social/professional role and identity* | Not important | 6 | 2 | 14 | 5 |  |
| **Increased self confidence** | Extremely important | 190 | 54 | 126 | 46 |  |
| *Beliefs about consequences* | Moderately important | 119 | 34 | 100 | 37 | 0.21 |
| *Goals* | Slightly important | 29 | 8 | 32 | 12 |  |
| *Emotion* | Not important | 14 | 4 | 14 | 5 |  |
| **Improved quality of life** | Extremely important | 235 | 67 | 134 | 49 |  |
| *Beliefs about consequences* | Moderately important | 99 | 28 | 90 | 33 | <0.001 |
| *Goals* | Slightly important | 17 | 5 | 38 | 14 |  |
| *Emotion* | Not important | 1 | <1 | 10 | 4 |  |
| **Improved social life** | Extremely important | 60 | 17 | 48 | 18 |  |
| *Beliefs about consequences* | Moderately important | 63 | 18 | 55 | 20 | 0.84 |
| *Goals* | Slightly important | 98 | 28 | 69 | 25 |  |
| *Social influences* |  |  |  |  |  |  |
| *Emotion* | Not important | 131 | 37 | 100 | 37 |  |
| **Improved pain management** | Extremely important | 82 | 23 | 43 | 16 |  |
| *Beliefs about consequences* | Moderately important | 84 | 24 | 52 | 19 | <0.001 |
| *Goals* | Slightly important | 72 | 21 | 48 | 18 |  |
| *Emotion* |  |  |  |  |  |  |
| *Reinforcement* | Not important | 114 | 32 | 129 | 47 |  |
| **Improved physical activity** | Extremely important | 89 | 25 | 61 | 22 |  |
| *Beliefs about consequences* | Moderately important | 147 | 42 | 103 | 38 | <0.001 |
| *Goals* | Slightly important | 90 | 26 | 59 | 22 |  |
| *Behavioural regulation* | Not important | 26 | 7 | 49 | 18 |  |
| **Improved diet quality** | Extremely important | 115 | 33 | 83 | 31 |  |
| *Beliefs about consequences* | Moderately important | 151 | 43 | 94 | 35 | **0.02** |
| *Goals* | Slightly important | 61 | 17 | 60 | 22 |  |
| *Behavioural regulation* | Not important | 25 | 7 | 35 | 13 |  |
| **Reduction in other medication** | Extremely important | 49 | 14 | 20 | 7 |  |
| *Beliefs about consequences* | Moderately important | 58 | 17 | 43 | 16 | 0.07 |
| *Goals* | Slightly important | 48 | 14 | 39 | 14 |  |
|  | Not important | 197 | 56 | 170 | 63 |  |

*TDF domains shown in italics*

### Discontinuation and post-treatment impact

All data related to discontinuation and post-treatment impact is presented in supplementary material 4 but summarised here. Former users identified cost of treatment (31%), planned end of treatment (25%) and side effects (25%) as the most prevalent reasons for stopping GLP-1RA medication. Similar responses were observed for barriers to restarting, with cost of treatment (33%), no longer requiring treatment (25%) and side effects (22%) most frequently reported. After stopping treatment, most participants reported weight gain (45%) or maintenance (40%), with 15% reporting weight loss. Pre-existing health conditions (60%), mental health (62%) and physical activity (58%) were typically unchanged after stopping treatment. Most participants reported an increase in the quantity of food consumed (56%), with the quality of food staying the same (52%).

### Subgroup analysis

Exploratory sub-group analyses were conducted in users across demographic factors (supplementary material 5-8). Statistically significant (p < 0.05) differences are summarised below by key themes but described in full in supplementary material 9.

### Access

Private online access was more common among females (current: 83% vs 57%; former: 68% vs 36%), younger (current: 80% vs 73%), and white users (current: 79% vs 61%; former: 62% vs 38%). Females self-weighed more (current: 83% vs 55%; former: 72% vs 39%), as did white participants (current: 80% vs 51%; former: 65% vs 40%). Current female users reported easier access (73% vs 63%), and white users greater comfort raising concerns (85% vs 70%). Higher socioeconomic status related to easier access in current users (74% vs 65%). Lower status former users used NHS face-to-face care more (30% vs 19%), and private services less (37% vs 47%). Former users with lower BMI at treatment accessed privately online (60% vs 52%) and self-weighed more (66% vs 49%). Former users with a higher BMI at treatment reported extremely challenging access more (17% vs 5%).

### Medication Experiences

Those that exercised reported increased physical activity whilst using medication (51% vs 30%). Among current users, lower BMI at survey was associated with greater improvements in quality of life (94% vs 81%), physical activity (65% vs 60%), mental health (63% vs 55%), and psychological support (33% vs 23%), while higher BMI users rated health outcomes (76% vs 57%) and weight loss (94% vs 74%) as more important. Among former users, lower BMI at survey related to improvements in quality of life (67% vs 43%), physical activity (52% vs 34%), and diet quality (75% vs 57%). Former male users more often reported increases in QoL (70% vs 52%), food quantity (38% vs 13%) and quality (46% vs 30%). Former users who exercise reported greater QoL (92% vs 76%) and mental-health (63% vs 47%) improvements.

### Healthcare Support

Current users with higher BMI at initiation more often reported inadequate dietary advice (28% vs 19%) and psychological support (50% vs 34%). Current users who exercised rated support more positively across provider knowledge (72% vs 54%), overall support (60% vs 40%), dietary advice (54% vs 34%), physical activity guidance (43% vs 22%), and psychological support (33% vs 16%). Former users with lower BMI at survey also rated support more positively: overall (62% vs 42%), dietary (56% vs 37%), physical activity (43% vs 28%), and psychological support (35% vs 18%). Former non-white users reported higher psychological support (55% vs 24%).

### Barriers to adherence

Current users with a higher BMI at treatment found cost extremely challenging more often (40% vs 25%). Among former users, lower socioeconomic status increased cost difficulties (42% vs 27%), and higher BMI at survey increased cost-related discontinuation (38% vs 14%). Former users with a higher BMI at survey also reported increased perceptions of cost (40% vs 29%) and side effects (32% vs 16%) as extremely challenging. Female former users cited cost (41% vs 13%) and side effects (44% vs 24%) more and discontinued for these reasons more (cost: 38% vs 14%; side effects: 31% vs 12%), while males more often stopped due to planned completion (50% vs 13%). Former users with higher BMI at survey reported greater discontinuation due to side effects (31% vs 12%) or cost (38% vs 14%).

### Motivators for adherence

Younger current users prioritised body image (76% vs 48%), self-confidence (65% vs 43%), and social life (26% vs 9%), while older users emphasised health improvements (74% vs 61%). Female current users rated weight loss (90% vs 78%), self-confidence (59% vs 40%), and pain management (27% vs 12%). Higher BMI at initiation was associated with placing more importance on health improvements (current only: 78% vs 61%), weight loss (current only: 93% vs 84%), and pain management (current: 28% vs 21%; former: 22% vs 14%). Non-white users rated social life (current: 32% vs 15%; former: 35% vs 15%) and medication reduction (current only: 27% vs 12%) more.

### Post-Discontinuation Outcomes

Female users reported more weight regain (52% vs 29%). Male users more frequently reported improvements in health conditions (36% vs 18%), physical activity (46% vs 28%), mental health (30% vs 18%), food quantity (38% vs 13%), and quality (46% vs 30%). Lower BMI at survey was associated with increased food intake (28% vs 8%) and improved diet quality (42% vs 24%). Non-white users increased food quantity (43% vs 17%). Those that exercised reported increased physical activity (39% vs 18%) and diet quality (38% vs 26%) more often post-discontinuation.

## Discussion

This study evaluated real-world experiences of GLP-1RA medications for obesity treatment among current and former users in the UK. Most participants reported that GLP-1RA treatment helped them achieve their weight goals, though substantial challenges emerged across access, healthcare support quality, and post-discontinuation outcomes, with disproportionate experiences across socio-demographic groups. Current users tended to report more positive treatment experiences and better support than former users, highlighting potential factors that may influence continuation. Encouragingly, post-discontinuation outcomes showed that 40% of former users maintained their weight and 15% reported further weight loss, though 45% experienced weight regain. These findings highlight the need for holistic, multidisciplinary care alongside GLP-1RA therapy and structured follow-up support to optimize long-term weight management outcomes beyond pharmacotherapy, with tailored approaches that address the distinct needs and barriers identified across different demographic groups.

Our findings demonstrate that most participants accessed GLP1-RA medication through private online services, with a smaller proportion accessing treatment via the NHS. This trend was more pronounced in current rather than former users, reflecting a potential shift towards a direct-to-consumer model of obesity care. Current users predominantly used Tirzepatide, likely reflecting superior weight loss efficacy [14] alongside market timing and availability. Most participants confirmed treatment eligibility through self-weighing with photographic verification, rather than in person clinical assessments. Whilst this method has demonstrated acceptable agreement with self-reported weight [15], it bypasses comprehensive clinical assessment. Cellular-connected scales may represent one strategy to verify self-reported weights and access objective health metrics. Younger and female current users were more likely to access medication privately online which may be attributable to a higher prevalence of body image dissatisfaction and appearance driven weight loss in these populations [16,17]. This may be more readily accommodated through less stringent eligibility criteria related to private online services. Former users with lower BMI at treatment initiation more frequently accessed medication privately and confirmed eligibility via self-weighing. Whilst this may partly be explained by NHS BMI eligibility thresholds, it may also suggest that those with psychosocial motivations may preferentially pursue private treatment, something which warrants investigation in future studies.

Participants generally reported adequate overall healthcare support during GLP-1RA treatment however perceived adequacy was consistently lower for physical activity, diet, and especially psychological care. This is concerning given clinical advice emphasises the integration of nutrition, physical activity and psychological support to maintain health and address potential mental health risks [18–20]. Current users reported more positive healthcare experiences than former users, suggesting that support quality may influence treatment continuation or alternatively that support structures are evolving as GLP1-RA medications become more embedded in clinical pathways. Former users from other ethnic groups reported notably better psychological support, however these findings should be interpreted with caution due to the small sample size of participants from minority ethnic backgrounds and the need to aggregate diverse groups into a single category for analysis. A concerning pattern emerged whereby those with the greatest clinical need reported the least adequate support. For example, current users with a higher BMI at treatment initiation reported experiencing worse diet and psychological support than those of a lower BMI. Similarly, current users not engaging in exercise, who may benefit most from behavioural guidance, rated support more negatively across all domains compared to those who do exercise. These disparities may contribute to differential treatment continuation and outcomes and provide strong recommendations for wrap around care.

Cost emerged as the most significant barrier to GLP-1RA use, with approximately one-third of current and former users rating it as extremely challenging. It was also identified as the primary reason for both discontinuation and barriers to restarting treatment in former users. This aligns with previous research [10] and is unsurprising given annual private costs for GLP-1RA medications in the UK range from approximately £2,000-£4,000 (UK pharmacy chains, October 2025). Notably, those with a lower self-defined social status faced disproportionate cost barriers, which is concerning given that obesity is inversely associated with socio-economic factors such as income [21]. The private model may therefore widen health inequalities by making treatment accessible primarily to those with financial means rather than clinical need [22]. Former female users and current users with a high BMI at the time of treatment also reported greater cost barriers, likely a result of preferential use of private services in women (68% vs. 36% in men), and longer treatment durations and therefore higher cumulative costs in those with a higher BMI.

Side effects were considerably more problematic for former users, with 21% rating them as extremely challenging compared to just 6% of current users. Consistent with previous research [10], side effects were one of the primary reasons for discontinuation (25%) and barrier to restarting (21%), suggesting tolerability may be a key determinant of treatment continuation. Although current and former users in this sample tended to use different medications, a recent clinical trial found no significant differences in rates of gastrointestinal symptoms between semaglutide and tirzepatide [14]. This suggests that the group differences observed here are unlikely to reflect inherent drug effects and instead may contribute to, or arise from, treatment discontinuation. The rates of discontinuation due to side effects reported in this study were higher than those typically observed in clinical trials, indicating a greater symptom burden under real-world conditions [23,24]. Optimisation of titration protocols (e.g. gradual, flexible titration over 16 weeks as opposed to 8 weeks) may represent an effective strategy to reduce side effects and facilitate treatment sustainability [25]. Former users with higher BMI at time of survey were more likely to cite side effects for discontinuation, suggesting a link between treatment intolerance and reduced efficacy or weight regain. Former female users also tended to identify side effects as a greater factor in discontinuation compared with males, consistent with previous evidence that females experience higher rates of gastrointestinal symptoms during GLP-1RA use [26,27].

The most reported motivator of adherence were weight loss, improved health outcomes, body image, self-confidence, and quality of life. Current users tended to rate these motivators as more important than former users, suggesting motivation may correlate with treatment continuation. Younger and female users prioritised psychosocial benefits (body image, self-confidence, social life), whereas older, male, higher BMI and non-white ethnic group users prioritised clinical outcomes (health markers, medication reduction, pain management). This aligns with previous research demonstrating that younger adults are more likely to be motivated by appearance and societal factors [28], despite being associated with poorer long-term weight loss outcomes compared to health-focused motivations [29,30]. The finding that higher BMI and non-white ethnic group users prioritised clinical outcomes likely reflects the greater burden of obesity-related complications and health disparities in these populations, highlighting the requirement for holistic, culturally sensitive care. Recognising diverse treatment motivations through personalised goal-setting and tailored support may be essential for optimising treatment outcomes in different patient populations.

Following discontinuation of GLP-1 receptor agonist therapy, around half of participants reported weight regain (45%), whilst 40% maintained weight (40%), and 15% lost further weight. As such, most users either therefore maintained or continued to lose weight post-discontinuation, which contrasts with typical weight gain trajectories in UK adults [31]. In those that did regain weight, this was likely largely explained by an increase in perceived food intake after stopping treatment. These findings somewhat align with recent evidence demonstrating that once GLP1-RA treatment is stopped, most or all of the weight is typically regained [32]. Importantly, those who engaged in regular exercise during treatment more frequently reported increases in physical activity and diet quality post-discontinuation. Given the known risks of sarcopenia associated with GLP-1RA-induced weight loss, this finding demonstrates the importance of supporting physical activity during treatment to facilitate sustained healthy behaviours and preserve muscle mass losses after discontinuation. Younger females reported more weight regain post-treatment, while males more often reported improvements in health, physical activity, and diet quality, suggesting better maintenance of healthy behaviours that may offset weight gain. The use of GLP-1RAs should be embedded within long-term, multidisciplinary care models that integrate behavioural, psychological and nutritional support to reduce the prevalence of weight regain and optimise health [20,33].

Despite providing valuable evidence on GLP-1RA user experiences in the UK, with key implications for policy and practice (Figure 1), several limitations should be considered. First, the cross-sectional design prevents causal inference and some associations may reflect reverse causality. Second, online survey tools (Prolific) may limit the demographic variation in the sample and limit generalisability to the UK population. For example, greater representation of individuals with lower socio-economic status and distinct minority ethnic groups warrants investigation. Third, we did not capture data on how long it had been since participants discontinued GLP1-RA treatment which somewhat limited interpretation of outcome data post-discontinuation. Finally, the use of self-reported and/or perceived outcomes, including changes in weight, may introduce social desirability and/or recall bias.

**Figure 1.**
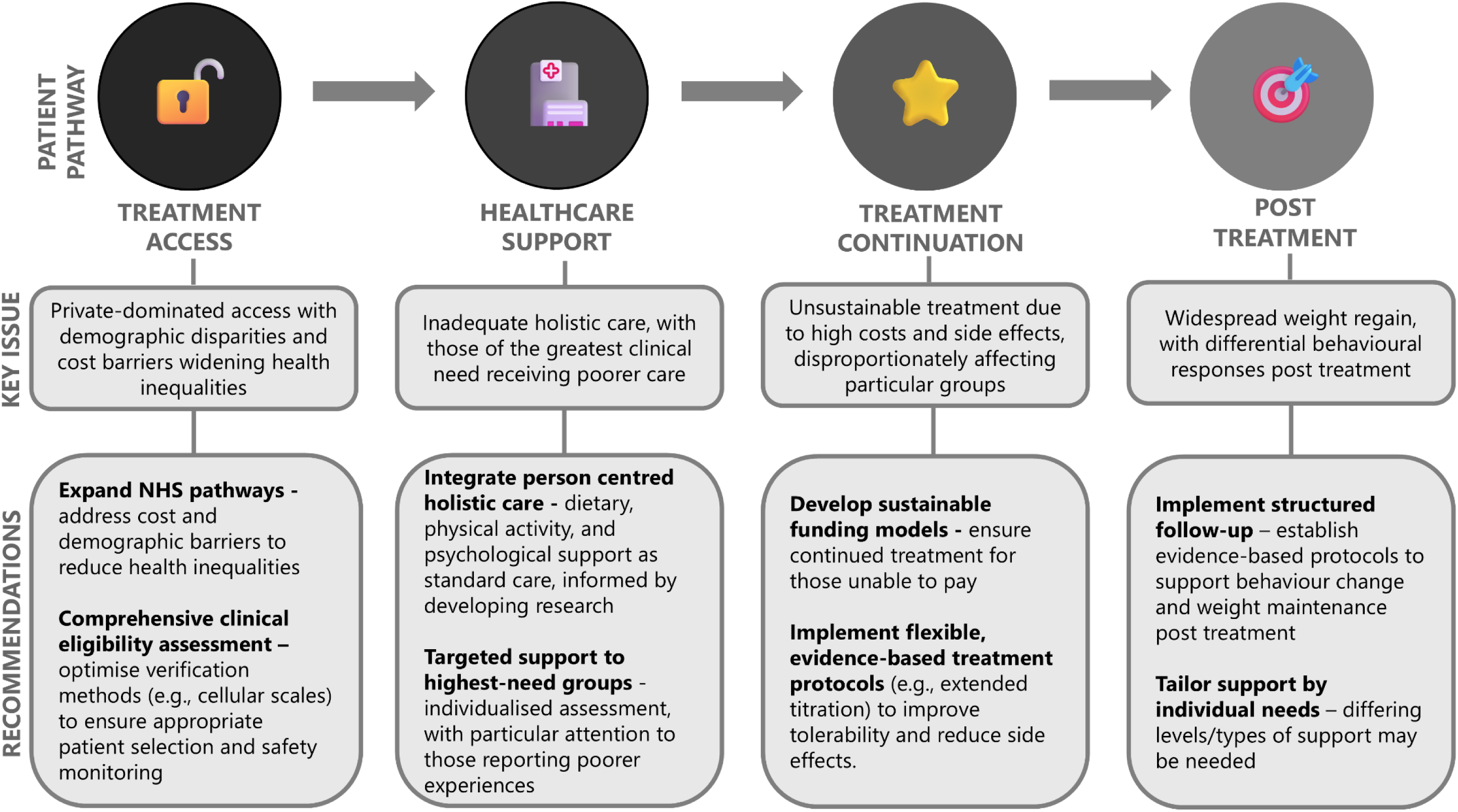
Summary of key findings, and policy and practice recommendations

In conclusion, whilst users generally reported that GLP-1RA treatment was effective, long-term usage is limited by a high cost, privately dominated access model, with inadequate holistic support. Former users tended to experience poorer healthcare support and side effects, which may provide insight into factors which explain treatment discontinuation. Future research should employ longitudinal designs to track the evolution of patient experiences, side effects, and weight trajectories. Future studies should also investigate the efficacy of tailored, multidisciplinary support models which are designed to address the distinct motivators and barriers identified in relevant subgroups.

## Acknowledgements

N/A

## Author contributions

AG and JM conceived the idea. AG, JM, OMS, KC and LE designed the questionnaire. AG conducted the analysis. All authors provided feedback on the findings and write up of the manuscript.

## Data Availability

The datasets generated during and/or analysed during the current study are available from the corresponding author on reasonable request.

