## Supplementary materials for "Exploring real-world user experiences of GLP-1 receptor agonist therapy for obesity treatment, and barriers and motivators to adherence"

**Supplementary material 1 - Full reporting of medication use, access and provision**

The type of GLP-1 receptor agonist used was not significantly different between former and current users ($p < 0.001$). A greater proportion of current users than former users were taking Tirzepatide (80% vs. 46%), while Semaglutide use was less common among current users (19% vs. 49%). Small proportions of current users reported using dulaglutide (<1%) and Liraglutide (<1%), and similarly small proportions of former users reporting using Liraglutide (3%), Exanatide (2%) or other (<1%) (which comprised answers of 'not sure'). The duration of GLP-1 receptor agonist medication use differed significantly between current and former users ($p = 0.04$). A lower percentage of current compared with former users took the medication for 0-6 months (57% vs. 66%), with smaller yet similar proportions reporting for 7-12 months (29% vs. 25%), 1-2 years (11% vs. 9%) or over two years (3% vs. <1%).

Dosage patterns are summarised for the two most frequently used drugs, Semaglutide and Tirzepatide:

- Starting dose: For those taking Tirzepatide, 94% of current and 82% of former users reported a starting dose of 2.5mg, with smaller proportions starting at 5mg (3% vs. 6%) or 7.5mg (both 1%). For Semaglutide, 69% of current and 62% of former users reported a start dose of 0.25mg, with smaller proportions reporting a higher start dose of 0.5-1.5mg (9% vs. 11%), 2-3mg (6% vs. 12%) or 5mg (5% vs. 5%).
- Current dose (current users only): Current Tirzepatide doses ranged from 2.5mg to 15mg, most commonly 5mg (28%), 7.5mg (29%) or 10mg (15%). Current Semaglutide doses ranged from 0.25mg to 25mg, most commonly 0.25-1mg (46%) or 2-2.5mg (18%).
- Maximum doses (former users only): For Tirzepatide, maximum doses typically clustered around 2.5mg (15%), 5mg (27%), 7.5 (16%), 10mg (16%) and 12-15mg (14%). For Semaglutide, they typically clustered around 0.25 (12%), 0.5 (12%), 1mg (23%), 2-3mg (17%), 5mg (9%) or 10mg (4%).

A greater proportion of current compared with former users reported increases in medication dosage (83% vs 68 %), whilst a lower proportion of current users reported dosage stayed the same (17% vs. 31%) ($p < 0.001$). Significant differences in how users accessed medication were observed (p <0.001). More current compared to formers users had their medication prescribed privately, mostly online (76% vs 58%) with some face to face (3% vs. 9%). A smaller proportion of current compared with former users accessed medication through an NHS healthcare provider online (4% vs. 7%) or face to face (16% vs. 23%). There were a small number of other responses which typically reported inclusion in clinical trials. There were significant differences between user groups when asked which healthcare provided manages their treatment ($p < 0.001$). A greater percentage of current compared with former users reported private healthcare provider (62% vs 44%), whilst less reported GP (16% vs. 22%) or none (11% vs. 23%). Similar percentages of users reported Specialist (6% vs 9%) and other (5% vs 2%) (which included pharmacies and institutions running clinical trials).

For both current and former users, 98% of participants had to confirm their weight prior to receiving their medication. A greater proportion of current users reported confirming their weight with self-weighing (76% vs. 61%), and less reporting being weighed by a professional (22% vs. 36%) ($p < 0.001$). Of those that self-weighed, a greater proportion of current compared with former users did have to send proof of weight in the form of photo or video (76% vs. 64%) ($p = 0.01$). A greater percentage of current compared with former users reported that their GP were aware they are taking GLP-1 medication (70% vs. 56%) ($p < 0.001$).

**Supplementary material 2 – Current and former user experiences of GLP-1 medication**

| Question | Response Options | Current users | | Former users | | *p* value |
|---|---|---|---|---|---|---|
| | | n | % | n | % | |
| **GLP1-RA medications are easy to use** | Agree | 337 | 96 | 253 | 93 | 0.2 |
| | Neutral | 10 | 3 | 10 | 4 | |
| | Disagree | 5 | 1 | 9 | 3 | |
| **GLP1-RA medications are easy to access** | Agree | 248 | 71 | 201 | 74 | 0.60 |
| | Neutral | 50 | 14 | 36 | 13 | |
| | Disagree | 54 | 15 | 35 | 13 | |
| **GLP-1 medications improved my overall quality of life** | Agree | 306 | 87 | 157 | 58 | **<0.001** |
| | Neutral | 39 | 11 | 58 | 21 | |
| | Disagree | 7 | 2 | 57 | 21 | |
| **I experienced significant side effects while using GLP1-RA medications** | Agree | 102 | 29 | 103 | 38 | **0.02** |
| | Neutral | 64 | 18 | 55 | 20 | |
| | Disagree | 186 | 53 | 114 | 42 | |
| **Side effects experienced** | Nausea | 206 | 59 | 166 | 61 | N/A |
| | Vomiting | 71 | 20 | 58 | 21 | |
| | Fatigue | 144 | 41 | 101 | 37 | |
| | Diarrhoea | 142 | 40 | 97 | 36 | |
| | Gallstones | 3 | 1 | 11 | 4 | |
| | Pancreatitis | 1 | <1 | 9 | 3 | |
| | Adverse psychological events | 9 | 3 | 13 | 5 | |
| | Constipation | 37 | 11 | 10 | 3.7 | |
| | Other | 34 | 10 | 30 | 11 | |
| | None | 34 | 10 | 28 | 10 | |
| **Since starting GLP1-RA medication, how has your physical activity level changed?** | Increased | 219 | 62 | 123 | 45 | **<0.001** |
| | Stayed the same | 118 | 34 | 118 | 43 | |
| | Decreased | 15 | 4 | 31 | 11 | |
| **Since starting GLP1-RAs, how has your** | Improved | 205 | 58 | 110 | 40 | **<0.001** |
| | Stayed the same | 137 | 39 | 130 | 48 | |

| | | | | | | |
|---|---|---|---|---|---|---|
| **mental health changed?** | Worsened | 10 | 3 | 32 | 12 | |
| **How has the quantity of food you consume been affected?** | Less food | 338 | 96 | 251 | 92 | **0.046** |
| | No change | 10 | 3 | 19 | 7 | |
| | More food | 4 | 1 | 2 | 1 | |
| **Do you feel GLP1-RA has improved the quality of your food choices?** | Healthier | 273 | 78 | 186 | 68 | **0.04** |
| | Stayed the same | 69 | 20 | 76 | 28 | |
| | Less healthy | 10 | 3 | 10 | 4 | |

**Supplementary material 3. Satisfaction and support perceptions of healthcare providers**

| Question | Response Options | Current users | | Former users | | *p* value |
|---|---|---|---|---|---|---|
| | | n | % | n | % | |
| **My healthcare provider is knowledgeable about GLP-1RA medication** | Agree | 261 | 74 | 182 | 67 | **<0.001** |
| | Neutral | 83 | 24 | 66 | 24 | |
| | Disagree | 8 | 2 | 24 | 9 | |
| **My healthcare provider provides adequate support during my treatment** | Agree | 217 | 62 | 148 | 54 | **0.02** |
| | Neutral | 103 | 29 | 80 | 29 | |
| | Disagree | 32 | 9 | 44 | 16 | |
| **My healthcare provider provides adequate dietary advice during my treatment** | Agree | 171 | 49 | 132 | 49 | 0.70 |
| | Neutral | 103 | 29 | 73 | 27 | |
| | Disagree | 78 | 22 | 67 | 25 | |
| **My healthcare provider provides adequate physical activity guidance during my treatment** | Agree | 127 | 36 | 101 | 37 | 0.71 |
| | Neutral | 99 | 28 | 82 | 30 | |
| | Disagree | 126 | 36 | 89 | 33 | |
| **My healthcare provider provides adequate psychological support** | Agree | 96 | 27 | 77 | 28 | 0.71 |
| | Neutral | 117 | 33 | 82 | 30 | |
| | Disagree | 139 | 40 | 113 | 42 | |
| **My healthcare provider regularly monitors my weight and health** | Agree | 216 | 61 | 141 | 52 | **0.048** |
| | Neutral | 49 | 14 | 52 | 19 | |
| | Disagree | 87 | 25 | 79 | 29 | |
| **I feel comfortable discussing my concerns with my healthcare provider** | Agree | 254 | 72 | 181 | 67 | 0.28 |
| | Neutral | 71 | 20 | 62 | 23 | |
| | Disagree | 27 | 8 | 28 | 10 | |

**Supplementary material 4. Discontinuation and post-treatment impact in former GLP-1 users**

| Question | Response options | n | % |
|---|---|---|---|
| **Reason for stopping GLP-1 medication** | Cost of treatment | 83 | 31 |
| | Planned end of treatment | 67 | 25 |
| | Side effects | 69 | 25 |
| | Lack of motivation/benefit | 12 | 4 |
| | Accessibility issues | 7 | 3 |
| | Stigma | 3 | 1 |
| | Changes to social life | 3 | 1 |
| | Other | 28 | 10 |
| **Barriers to restarting GLP-1 medication** | Cost of treatment | 90 | 33 |
| | Planned end of treatment | 69 | 25 |
| | Side effects | 59 | 22 |
| | Lack of motivation/benefit | 26 | 10 |
| | Accessibility issues | 11 | 4 |
| | Stigma | 0 | 0 |
| | Changes to social life | 2 | 1 |
| | Other | 15 | 6 |
| **What happened to your weight after you stopped taking GLP-1 medication?** | Gained weight | 123 | 45 |
| | Maintained | 110 | 40 |
| | Lost weight | 39 | 15 |
| **What happened to your other health conditions after you stopped taking GLP-1 medication?** | Improvement | 39 | 24 |
| | No change | 106 | 66 |

| | | | |
|---|---|---|---|
| | Worsening | 15 | 9 |
| **What happened to your physical activity after you stopped taking GLP-1 medication?** | Increased | 91 | 34 |
| | No change | 158 | 58 |
| | Decreased | 23 | 9 |
| **What happened to your mental health after you stopped taking GLP-1 medication?** | Improvement | 59 | 22 |
| | No change | 168 | 62 |
| | Worsening | 45 | 17 |
| **What happened to the quantity of food you consumed when you stopped taking GLP-1 medication?** | Increased | 56 | 21 |
| | No change | 63 | 23 |
| | Decreased | 153 | 56 |
| **What happened to the quality of food you consumed when you stopped taking GLP-1 medication?** | Increased | 95 | 35 |
| | No change | 140 | 52 |
| | Decreased | 37 | 14 |

**Supplementary material 5. Subgroup analysis of current GLP-1 user experiences, and barriers and motivators to adherence by age, gender and BMI.**

| Question | Response Options | Age | | | Gender | | | BMI at time of survey | | | BMI at start of treatment | | |
|---|---|---|---|---|---|---|---|---|---|---|---|---|---|
| | | Youn-ger | Older | *p* | Male | Female | *p* | ≤ 30 | >30 | *p* | 25-39.9 | 40+ | *p* |
| **Medication use, access and provision** | | | | | | | | | | | | | |
| Type of drug | Semaglutide | 35 (20) | 32 (18) | 0.52 | 20 (24) | 46 (17) | 0.47 | 30 (19) | 37 (19) | 0.63 | 39 (17) | 28 (23) | 0.14 |
| | Tirzepatide | 137 (80) | 146 (81) | | 63 (76) | 220 (82) | | 131 (81) | 152 (80) | | 189 (83) | 94 (76) | |
| | Dulaglutide | 0 (0) | 1 (0.6) | | 0 (0) | 1 (<1) | | 0 (0) | 1 (<1) | | 0 (0) | 1 (1) | |
| | Liraglutide | 0 (0) | 1 (0.6) | | 0 (0) | 1 (<1) | | 0 (0) | 1 (<1) | | 0 (0) | 1 (1) | |
| Duration of use | 0-6 months | 87 (51) | 114 (63) | **0.02** | 48 (58) | 153 (57) | 0.17 | 86 (53) | 115 (60) | 0.43 | 72 (58) | 72 (58) | 0.90 |
| | 7-12 months | 53 (31) | 50 (28) | | 19 (23) | 84 (31) | | 53 (33) | 50 (26) | | 69 (30) | 34 (27) | |
| | 1-2 years | 25 (15) | 14 (8) | | 12 (15) | 26 (10) | | 19 (12) | 20 (11) | | 25 (11) | 14 (11) | |
| | 2 + years | 7 (4) | 2 (1) | | 4 (5) | 5 (2) | | 3 (2) | 6 (3) | | 5 (2) | 4 (3) | |
| Access to medication | Privately online | 137 (80) | 132 (73) | **0.04** | 47 (57) | 222 (83) | **<0.001** | 127 (79) | 142 (74) | 0.12 | 178 (78) | 91 (73) | 0.24 |
| | Privately face to face | 5 (3) | 2 (3) | | 4 (5) | 6 (2) | | 6 (4) | 4 (2) | | 7 (3) | 3 (2) | |
| | NHS provider online | 9 (5) | 4 (2) | | 4 (5) | 9 (3) | | 3 (2) | 10 (5) | | 7 (3) | 6 (5) | |
| | NHS provider face to face | 21 (12) | 34 (19) | | 26 (31) | 28 (10) | | 21 (13) | 34 (18) | | 31 (14) | 24 (19) | |

| | | | | | | | | | | | | | |
|---|---|---|---|---|---|---|---|---|---|---|---|---|---|
| | Other | 0 (0) | 5 (3) | | 2 (2) | 3 (1)) | | 4 (3) | 1 (1) | | 5 (2) | 0 (0) | |
| Which healthcare provider manages your treatment | Private healthcare | 106 (62) | 112 (62) | 0.06 | 37 (45) | 181 (68) | **<0.001** | 103 (64) | 115 (60) | 0.64 | 139 (61) | 79 (64) | 0.57 |
| | GP | 22 (13) | 33 (18) | | 26 (31) | 29 (11) | | 21 (13) | 34 (18) | | 35 (15) | 20 (16) | |
| | Specialist | 13 (8) | 7 (4) | | 10 (12) | 9 (3) | | 10 (6) | 10 (5) | | 11 (5) | 9 (7) | |
| | None | 25 (15) | 15 (8) | | 6 (7) | 34 (13) | | 20 (12) | 20 (11) | | 30 (13) | 10 (8) | |
| | Other | 6 (4) | 13 (7) | | 4 (5) | 15 (6) | | 7 (4) | 12 (6) | | 13 (6) | 6 (5) | |
| Method of confirming weight | Self-weighed | 132 (77) | 137 (76) | 0.88 | 46 (55) | 223 (83) | **<0.001** | 124 (77) | 145 (76) | 0.76 | 177 (78) | 92 (74) | 0.29 |
| | Weighed by professional | 36 (21) | 40 (22) | | 36 (43) | 39 (15) | | 33 (21) | 43 (23) | | 45 (20) | 31 (25) | |
| | None | 4 (2) | 3 (2) | | 1 (1) | 6 (2) | | 4 (3) | 3 (2) | | 6 (3) | 1 (1) | |
| If self-weighed, did you have to send proof of weight | Yes | 99 (75) | 104 (76) | 0.86 | 32 (70) | 171 (77) | 0.31 | 92 (74) | 32 (26) | 0.65 | 139 (79) | 64 (70) | 0.11 |
| | No | 33 (25) | 33 (24) | | 14 (30) | 52 (23) | | 111 (77) | 34 (23) | | 38 (22) | 28 (30) | |
| **Experiences of using GLP-1 medication** | | | | | | | | | | | | | |
| GLP1-RA medications are easy to use | Agree | 165 (96) | 172 (96) | 0.75 | 80 (96) | 256 (96) | 0.41 | 154 (96) | 183 (96) | 0.93 | 217 (95) | 120 (97) | 0.72 |
| | Neutral | 4 (2) | 6 (3) | | 1 (1) | 9 (3) | | 5 (3) | 5 (3) | | 7 (3) | 3 (2) | |
| | Disagree | 3 (2) | 2 (1) | | 2 (2) | 3 (1) | | 2 (1) | 3 (2) | | 4 (2) | 1 (1) | |

| | | | | | | | | | | | | | |
|---|---|---|---|---|---|---|---|---|---|---|---|---|---|
| GLP1-RA medications are easy to access | Agree | 115 (67) | 133 (74) | 0.23 | 52 (63) | 196 (73) | **0.03** | 118 (73) | 130 (68) | 0.06 | 164 (72) | 84 (68) | 0.47 |
| | Neutral | 25 (15) | 25 (14) | | 19 (23) | 30 (11) | | 26 (16) | 24 (13) | | 33 (15) | 17 (14) | |
| | Disagree | 32 (19) | 22 (12) | | 12 (15) | 42 (16) | | 17 (11) | 37 (19) | | 31 (14) | 23 (19) | |
| GLP-1 medications improved my overall quality of life | Agree | 146 (85) | 160 (89) | 0.54 | 72 (87) | 233 (87) | 0.81 | 151 (94) | 155 (81) | **<0.001** | 204 (90) | 102 (82) | 0.08 |
| | Neutral | 22 (13) | 17 (9) | | 10 (12) | 29 (11) | | 9 (6) | 30 (16) | | 19 (8) | 20 (16) | |
| | Disagree | 4 (2) | 3 (2) | | 1 (1) | 6 (2) | | 1 (<1) | 6 (3) | | 5 (2) | 2 (2) | |
| I experienced significant side effects while using GLP1-RA medications | Agree | 56 (33) | 46 (26) | 0.35 | 27 (33) | 75 (28) | 0.31 | 52 (32) | 50 (26) | 0.45 | 69 (30) | 33 (27) | 0.68 |
| | Neutral | 30 (17) | 34 (19) | | 18 (22) | 45 (17) | | 28 (17) | 36 (19) | | 39 (17) | 25 (20) | |
| | Disagree | 86 (50) | 100 (56) | | 38 (46) | 148 (55) | | 81 (50) | 105 (55) | | 120 (53) | 66 (53) | |
| Since starting GLP1-RA medication, how has your physical activity level changed? | Increased | 108 (63) | 111 (62) | 0.11 | 58 (70) | 160 (60) | 0.25 | 105 (65) | 114 (60) | **0.02** | 138 (61) | 81 (65) | 0.18 |
| | Stayed the same | 53 (31) | 65 (36) | | 22 (27) | 96 (36) | | 45 (28) | 73 (38) | | 77 (34) | 41 (33) | |
| | Decreased | 11 (6) | 4 (2) | | 3 (4) | 12 (5) | | 11 (7) | 4 (2) | | 13 (6) | 2 (2) | |
| Since starting GLP1-RAs, how has your mental health changed? | Improved | 100 (58) | 105 (58) | 0.13 | 42 (51) | 162 (60) | 0.07 | 101 (63) | 104 (55) | **0.046** | 132 (58) | 73 (59) | 0.92 |
| | Stayed the same | 64 (37) | 73 (41) | | 36 (43) | 101 (38) | | 53 (33) | 84 (44) | | 90 (40) | 47 (38) | |

| | | | | | | | | | | | | | |
|---|---|---|---|---|---|---|---|---|---|---|---|---|---|
| | Worsened | 8 (5) | 2 (1) | | 5 (6) | 5 (6) | | 7 (4) | 3 (2) | | 6 (3) | 4 (3) | |
| How has the quantity of food you consume been affected? | Less food | 163 (95) | 175 (97) | 0.12 | 78 (94) | 259 (97) | 0.47 | 155 (96) | 183 (96) | 0.30 | 217 (95) | 121 (97) | 0.31 |
| | No change | 5 (3) | 5 (3) | | 4 (5) | 6 (2) | | 3 (2) | 7 (4) | | 7 (3) | 3 (2) | |
| | More food | 4 (2) | 0 (0) | | 1 (1) | 3 (1) | | 3 (2) | 1 (<1) | | 4 (2) | 0 (0) | |
| Do you feel GLP1-RA has improved the quality of your food choices? | Healthier | 131 (76) | 142 (79) | 0.71 | 67 (81) | 205 (77) | 0.54 | 121 (75) | 152 (80) | 0.50 | 176 (77) | 97 (78) | 0.59 |
| | Stayed the same | 35 (20) | 34 (19) | | 13 (16) | 56 (21) | | 34 (21) | 35 (18) | | 44 (19) | 25 (20) | |
| | Less healthy | 6 (4) | 4 (2) | | 3 (4) | 7 (3) | | 6 (4) | 4 (2) | | 8 (4) | 2 (2) | |
| **Healthcare provider support** | | | | | | | | | | | | | |
| My healthcare provider is knowledgeable about GLP-1RA medication | Agree | 128 (74) | 133 (74) | 0.70 | 67 (81) | 193 (72) | 0.14 | 128 (80) | 133 (70) | 0.09 | 172 (75) | 89 (72) | 0.58 |
| | Neutral | 39 (23) | 44 (24) | | 16 (19) | 67 (25) | | 31 (19) | 52 (27) | | 52 (23) | 31 (25) | |
| | Disagree | 5 (3) | 3 (2) | | 0 (0) | 8 (3) | | 2 (1) | 6 (3) | | 4 (2) | 4 (3) | |
| My healthcare provider provides adequate support | Agree | 103 (60) | 114 (63) | 0.64 | 57 (69) | 159 (59) | 0.29 | 107 (67) | 110 (58) | 0.07 | 147 (65) | 70 (57) | 0.22 |
| | Neutral | 51 (30) | 52 (29) | | 19 (23) | 84 (31) | | 45 (28) | 58 (30) | | 64 (28) | 39 (32) | |
| | Disagree | 18 (10) | 14 (8) | | 7 (8) | 25 (9) | | 9 (6) | 23 (12) | | 17 (8) | 15 (12) | |

| | | | | | | | | | | | | | |
|---|---|---|---|---|---|---|---|---|---|---|---|---|---|
| My healthcare provider provides adequate dietary advice during my treatment | Agree | 87 (51) | 84 (47) | 0.46 | 48 (58) | 122 (46) | 0.14 | 85 (53) | 86 (45) | 0.13 | 109 (48) | 62 (50) | **0.03** |
| | Neutral | 45 (26) | 58 (32) | | 21 (25) | 82 (31) | | 48 (30) | 55 (29) | | 76 (33) | 27 (22) | |
| | Disagree | 40 (23) | 38 (21) | | 14 (17) | 64 (24) | | 28 (17) | 50 (26) | | 43 (19) | 35 (28) | |
| My healthcare provider provides adequate physical activity guidance | Agree | 66 (38) | 61 (34) | 0.67 | 38 (46) | 88 (33) | 0.10 | 68 (42) | 59 (31) | 0.08 | 88 (39) | 39 (32) | 0.13 |
| | Neutral | 46 (27) | 53 (29) | | 21 (25) | 78 (29) | | 43 (27) | 56 (29) | | 67 (29) | 32 (26) | |
| | Disagree | 60 (35) | 66 (37) | | 24 (29) | 24 (29) | | 50 (31) | 76 (40 | | 73 (32) | 53 (43) | |
| My healthcare provider provides adequate psychological support | Agree | 49 (29) | 47 (26) | 0.76 | 28 (34) | 67 (25) | 0.07 | 53 (33) | 43 (23) | **0.01** | 68 (30) | 28 (23) | **0.01** |
| | Neutral | 54 (31) | 63 (35) | | 31 (37) | 86 (32) | | 57 (35) | 60 (31) | | 83 (36) | 34 (27) | |
| | Disagree | 69 (40) | 70 (39) | | 24 (29) | 115 (43) | | 52 (32) | 88 (46) | | 77 (34) | 62 (50) | |
| My healthcare provider regularly monitors my weight and health | Agree | 99 (58) | 117 (65) | 0.26 | 50 (60) | 165 (62) | 0.42 | 105 (65) | 111 (58) | 0.36 | 132 (59) | 82 (66) | 0.40 |
| | Neutral | 24 (14) | 25 (14) | | 15 (18) | 34 (13) | | 19 (12) | 30 (16) | | 34 (15) | 15 (12) | |
| | Disagree | 49 (29) | 38 (21) | | 18 (22) | 69 (26) | | 37 (23) | 50 (26) | | 60 (26) | 27 (22) | |
| I feel comfortable discussing my concerns with my healthcare provider | Agree | 121 (70) | 133 (74) | 0.76 | 64 (77) | 189 (71) | 0.33 | 120 (75) | 134 (70) | 0.38 | 164 (72) | 90 (73) | 0.98 |
| | Neutral | 37 (22) | 34 (19) | | 12 (15) | 59 (22) | | 32 (20) | 39 (20) | | 46 (20) | 25 (20) | |

| | | | | | | | | | | | | | |
|---|---|---|---|---|---|---|---|---|---|---|---|---|---|
| | Disagree | 14 (8) | 13 (7) | | 7 (8) | 20 (8) | | 9 (6) | 18 (9)- | | 18 (8) | 9 (7) | |
| **Barriers to adherence** | | | | | | | | | | | | | |
| **Cost of medication** | Extremely challenging | 54 (31) | 53 (29) | **0.053** | 20 (24) | 86 (32) | 0.11 | 42 (26) | 65 (34) | 0.18 | 58 (25) | 49 (40) | **0.02** |
| | Moderately challenging | 69 (40) | 62 (34) | | 27 (33) | 104 (39) | | 63 (39) | 68 (36) | | 91 (40) | 40 (32) | |
| | Slightly challenging | 28 (16) | 23 (13) | | 17 (21) | 34 (13) | | 29 (18) | 22 (12) | | 39 (17) | 12 (10) | |
| | Not challenging | 21 (12) | 42 (23) | | 19 (23) | 44 (16) | | 27 (17) | 36 (19) | | 40 (18) | 23 (19) | |
| **Side effects** | Extremely challenging | 12 (7) | 8 (4) | 0.52 | 4 (5) | 16 (6) | 0.85 | 9 (6) | 11 (6) | 0.98 | 13 (6) | 7 (6) | 0.74 |
| | Moderately challenging | 32 (19) | 31 (17) | | 17 (21) | 45 (17) | | 29 (18) | 34 (18) | | 37 (16) | 26 (21) | |
| | Slightly challenging | 64 (37) | 62 (34) | | 28 (34) | 98 (37) | | 56 (35) | 70 (37) | | 84 (37) | 42 (34) | |
| | Not challenging | 64 (37) | 79 (44) | | 34 (41) | 109 (41) | | 67 (42) | 76 (40) | | 94 (41) | 49 (40) | |
| **Access to medication** | Extremely challenging | 9 (5) | 4 (2) | 0.07 | 6 (7) | 7 (3) | 0.13 | 6 (4) | 7 (4) | 0.99 | 8 (4) | 5 (4) | 0.92 |
| | Moderately challenging | 24 (14) | 28 (16) | | 14 (7) | 37 (14) | | 23 (14) | 29 (15) | | 33 (15) | 19 (15) | |
| | Slightly challenging | 51 (30) | 37 (21) | | 23 (28) | 65 (24) | | 40 (25) | 48 (25) | | 55 (24) | 33 (27) | |
| | Not challenging | 88 (51) | 111 (62) | | 40 (48) | 159 (59) | | 92 (57) | 107 (56) | | 132 (58) | 67 (54) | |
| **Social stigma** | Extremely challenging | 15 (9) | 12 (7) | 0.32 | 5 (6) | 21 (8) | 0.70 | 10 (6) | 17 (9) | 0.27 | 16 (7) | 11 (9) | 0.38 |
| | Moderately challenging | 37 (22) | 31 (17) | | 13 (16) | 55 (21) | | 26 (16) | 42 (22) | | 39 (17) | 29 (23) | |

| | | | | | | | | | | | | | |
|---|---|---|---|---|---|---|---|---|---|---|---|---|---|
| | Slightly challenging | 46 (27) | 42 (23) | | 22 (27) | 66 (25) | | 46 (29) | 42 (22) | | 57 (25) | 31 (25) | |
| | Not challenging | 74 (43) | 95 (53) | | 43 (52) | 126 (47) | | 79 (49) | 90 (47) | | 116 (51) | 53 (43) | |
| **Negative effects on social life** | Extremely challenging | 5 (3) | 8 (4) | **<0.01** | 2 (2) | 11 (4) | 0.59 | 7 (4) | 6 (3) | 0.90 | 7 (3) | 6 (5) | 0.75 |
| | Moderately challenging | 33 (19) | 13 (7) | | 10 (12) | 35 (13) | | 22 (14) | 24 (13) | | 29 (13) | 17 (14) | |
| | Slightly challenging | 53 (31) | 50 (28) | | 21 (25) | 82 (31) | | 45 (28) | 58 (30) | | 65 (29) | 38 (31) | |
| | Not challenging | 81 (47) | 109 (61) | | 50 (60) | 140 (52) | | 87 (54) | 103 (54) | | 127 (56) | 63 (51) | |
| **Motivators to adherence** | | | | | | | | | | | | | |
| **Improved health outcomes** | Extremely important | 104 (61) | 133 (74) | **0.02** | 53 (64) | 184 (69) | 0.38 | 92 (57) | 145 (76) | **<0.01** | 140 (61) | 97 (78) | **0.01** |
| | Moderately important | 60 (35) | 41 (23) | | 25 (30) | 75 (28) | | 61 (38) | 40 (21) | | 77 (34) | 24 (19) | |
| | Slightly important | 8 (5) | 4 (2) | | 5 (6) | 7 (3) | | 7 (4) | 5 (3) | | 10 (4) | 2 (2) | |
| | Not important | 0 (0) | 2 (1 | | 0 (0) | 2 (1) | | 1 (<1) | 1 (<1) | | 1 (<1) | 1 (1) | |
| **Weight loss** | Extremely important | 146 (85) | 160 (89) | 0.38 | 65 (78) | 241 (90) | **<0.01** | 127 (79) | 179 (94) | **<0.001** | 191 (84) | 115 (93) | **0.051** |
| | Moderately important | 25 (15) | 18 (10) | | 18 (22) | 24 (9) | | 33 (21) | 10 (5) | | 35 (15) | 8 (7) | |
| | Slightly important | 1 (1) | 2 (1) | | 0 (0) | 3 (1) | | 1 (<1) | 2 (1) | | 2 (1) | 1 91) | |
| | Not important | 0 (0) | 0 (0) | | 0 (0) | 0 (0) | | 0 (0) | 0 (0) | | 0 (0) | 0 (0) | |
| **Improved body image** | Extremely important | 130 (76) | 87 (48) | **<0.001** | 45 (54) | 172 (62) | 0.26 | 103 (64) | 114 (60) | 0.73 | 147 (65) | 70 (57) | 0.39 |

| | | | | | | | | | | | | | |
|---|---|---|---|---|---|---|---|---|---|---|---|---|---|
| | Moderately important | 29 (17) | 69 (38) | | 25 (30) | 72 (27) | | 44 (27) | 54 (28) | | 61 (27) | 37 (30) | |
| | Slightly important | 11 (6) | 20 (11) | | 11 (13) | 20 (8) | | 12 (8) | 19 (10) | | 17 (8) | 14 (11) | |
| | Not important | 2 (1) | 4 (2) | | 2 (2) | 4 (2) | | 2 (1) | 4 (2) | | 3 (1) | 3 (2) | |
| **Increased self confidence** | Extremely important | 112 (65) | 78 (43) | **<0.001** | 33 (40) | 157 (59) | **0.03** | 83 (52) | 107 (56) | 0.29 | 126 (55) | 64 (52) | 0.41 |
| | Moderately important | 49 (29) | 70 (39) | | 36 (43) | 82 (31) | | 62 (39) | 57 (30) | | 79 (35) | 40 (32) | |
| | Slightly important | 9 (5) | 20 (11) | | 9 (11) | 20 (8) | | 10 (6) | 19 (10) | | 15 (7) | 14 (11) | |
| | Not important | 2 (1) | 12 (7) | | 5 (6) | 9 (3) | | 6 (4) | 8 (4) | | 8 (4) | 6 (5) | |
| **Improved quality of life** | Extremely important | 122 (71) | 113 (63) | 0.33 | 47 (57) | 187 (70) | 0.13 | 104 (65) | 131 (69) | 0.60 | 153 (67) | 82 (66) | 0.80 |
| | Moderately important | 43 (25) | 56 (31) | | 30 (36) | 69 (26) | | 47 (29) | 52 (27) | | 62 (27) | 37 (30) | |
| | Slightly important | 7 (4) | 10 (6) | | 6 (7) | 11 (4) | | 9 (6) | 8 (4) | | 12 (5) | 5 (4) | |
| | Not important | 0 (0) | 1 (1) | | 0 (0) | 1 (<1) | | 1 (<1) | 0 (0) | | 1 (<1) | 0 (0) | |
| **Improved social life** | Extremely important | 44 (26) | 16 (9) | **<0.001** | 12 (15) | 47 (18) | 0.78 | 28 (17) | 32 (17) | 0.65 | 36 (16) | 24 (19) | 0.81 |
| | Moderately important | 33 (19) | 30 (17) | | 16 (19) | 47 (18) | | 33 (21) | 30 (16) | | 40 (18) | 23 (19) | |
| | Slightly important | 40 (23) | 58 (32) | | 26 (31) | 72 (27) | | 44 (27) | 54 (28) | | 66 (29) | 32 (16) | |
| | Not important | 55 (32) | 76 (42) | | 29 (35) | 102 (38) | | 56 (35) | 75 (39) | | 86 (38) | 45 (36) | |
| **Improved pain management** | Extremely important | 38 (22) | 44 (24) | 0.69 | 10 (12) | 71 (27) | **<0.01** | 32 (20) | 50 (26) | 0.27 | 47 (21) | 35 (28) | **<0.001** |

| | | | | | | | | | | | | | |
|---|---|---|---|---|---|---|---|---|---|---|---|---|---|
| | Moderately important | 39 (23) | 45 (25) | | 20 (24) | 64 (24) | | 36 (22) | 48 (25) | | 45 (20) | 39 (32) | |
| | Slightly important | 34 (20) | 38 (21) | | 15 (18) | 57 (21) | | 33 (21) | 39 (20) | | 48 (21) | 24 (19) | |
| | Not important | 61 (36) | 53 (29) | | 38 (46) | 76 (28) | | 60 (37) | 54 (28) | | 88 (39) | 26 (21) | |
| **Improved physical activity** | Extremely important | 46 (27) | 43 (24) | 0.47 | 12 (15) | 76 (28) | **0.048** | 38 (24) | 51 (17) | 0.34 | 56 (25) | 33 (27) | 0.13 |
| | Moderately important | 68 (40) | 79 (44) | | 43 (52) | 104 (39) | | 64 (40) | 83 (44) | | 90 (40) | 57 (46) | |
| | Slightly important | 48 (28) | 42 (23) | | 23 (28) | 67 (25) | | 43 (27) | 47 (25) | | 60 (26) | 30 (24) | |
| | Not important | 10 (6) | 16 (9) | | 5 (6) | 21 (8) | | 16 (10) | 10 (5) | | 22 (10) | 4 (3) | |
| **Improved diet quality** | Extremely important | 57 (33) | 58 (32) | 0.40 | 21 (25) | 93 (35) | 0.44 | 51 (32) | 64 (34) | 0.12 | 71 (31) | 44 (36) | **0.03** |
| | Moderately important | 79 (46) | 72 (40) | | 40 (48) | 111 (41) | | 62 (39) | 89 (47) | | 90 (40) | 61 (49) | |
| | Slightly important | 24 (14) | 37 (21) | | 15 (18) | 46 (17) | | 32 (20) | 29 (15) | | 47 (21) | 14 (11) | |
| | Not important | 12 (7) | 13 (7) | | 7 (8) | 18 (7) | | 16 (10) | 9 (5) | | 20 (9) | 5 (4) | |
| **Reduction in other medication** | Extremely important | 17 (10) | 32 (18) | 0.06 | 10 (12) | 39 (15) | 0.72 | 21 (13) | 28 (15) | 0.93 | 30 (13) | 19 (15) | 0.15 |
| | Moderately important | 26 (15) | 32 (18) | | 11 (13) | 46 (17) | | 26 (16) | 32 (17) | | 37 (16) | 21 (17) | |
| | Slightly important | 21 (12) | 27 (15) | | 13 (16) | 35 (13) | | 21 (13) | 27 (14) | | 25 (11) | 23 (19) | |
| | Not important | 108 (63) | 89 (49) | | 49 (59) | 148 (55) | | 93 (58) | 104 (55) | | 136 (60) | 61 (49) | |

**Supplementary material 6. Subgroup analysis of current GLP-1 user experiences, and barriers and motivators to adherence by ethnicity, education, social status and physical activity.**

| Question | Response Options | Ethnicity | | | Education | | | Social status | | | Physical activity | | |
|---|---|---|---|---|---|---|---|---|---|---|---|---|---|
| | | White | Other | *p* | Lower | Higher | *p* | Lower | Higher | *p* | No | Yes | *p* |
| **Medication use, access and provision** | | | | | | | | | | | | | |
| Type of drug | Semaglutide | 57 (18) | 10 (24) | **0.03** | 19 (15) | 48 (21) | 0.38 | 26 (19) | 41 (19) | 0.72 | 16 (15) | 51 (21) | 0.38 |
| | Tirzepatide | 253 (81) | 30 (73) | | 106 (85) | 177 (78) | | 113 (81) | 170 (80) | | 94 (86) | 189 (78) | |
| | Dulaglutide | 0 (0) | 1 (2) | | 0 (0) | 1 (<1) | | 0 (0) | 1 (1) | | 0 (0) | 1 (<1) | |
| | Liraglutide | 1 (0) | 0 (0) | | 0 (0) | 1 (<1) | | 0 (0) | 1 (1) | | 0 (0) | 1 (<1) | |
| Duration of use | 0-6 months | 175 (56) | 26 (63) | 0.75 | 77 (62) | 124 (55) | 0.48 | 78 (56) | 123 (58) | 0.40 | 67 (61) | 134 (55) | 0.77 |
| | 7-12 months | 94 (30) | 9 (22) | | 35 (28) | 68 (30) | | 37 (27) | 66 (31) | | 30 (27) | 73 (30) | |
| | 1-2 years | 34 (11) | 5 (12) | | 10 (8) | 29 (13) | | 20 (14) | 19 (9) | | 11 (10) | 28 (12) | |
| | 2 + years | 8 (3) | 1 (2) | | 3 (2) | 6 (3) | | 4 (3) | 5 (2) | | 2 (2) | 7 (3) | |
| Access to medication | Privately online | 244 (79) | 25 (61) | **0.054** | 100 (80) | 169 (74) | 0.47 | 104 (75) | 165 (78) | 0.57 | 78 (71) | 191 (79) | **0.02** |
| | Privately face to face | 7 (2) | 3 (7) | | 3 (2) | 7 (3) | | 4 (3) | 6 (3) | | 0 (0) | 10 (4) | |
| | NHS provider online | 11 (4) | 2 (5) | | 6 (5) | 7 (3) | | 3 (2) | 10 (5) | | 7 (6) | 6 (3) | |
| | NHS provider face to face | 44 (14) | 11 (27) | | 14 (11) | 41 (18) | | 26 (19) | 29 (14) | | 23 (21) | 32 (13) | |

| | | | | | | | | | | | | |
|---|---|---|---|---|---|---|---|---|---|---|---|---|
| | Other | 5 (2) | 0 (0) | | 2 (2) | 3 (1) | | 2 (1) | 3 (1) | | 2 (2) | 3 (1) | |
| Which healthcare provider manages your treatment | Private healthcare | 197 (63) | 21 (51) | 0.16 | 79 (63) | 139 (61) | 0.63 | 81 (58) | 137 (64) | 0.73 | 61 (56) | 61 (56) | **0.02** |
| | GP | 44 (14) | 44 (14) | | 19 (15) | 36 (16) | | 24 (17) | 31 (15) | | 26 (24) | 29 (12) | |
| | Specialist | 15 (5) | 16 (5) | | 4 (3) | 16 (7) | | 10 (7) | 10 (5) | | 6 (6) | 14 (6) | |
| | None | 37 (12) | 37 (12) | | 15 (12) | 25 (11) | | 17 (12) | 23 (11) | | 15 (14) | 25 (10) | |
| | Other | 17 (6) | 17 (6) | | 8 (6) | 11 (5) | | 7 (5) | 12 (6) | | 2 (2) | 17 (7) | |
| Method of confirming weight | Self-weighed | 248 (80) | 21 (51) | **<0.001** | 106 (85) | 163 (72) | **0.02** | 107 (77) | 162 (76) | 0.84 | 80 (73) | 189 (78) | 0.23 |
| | Weighed by professional | 58 (19) | 19 (44) | | 18 (14) | 58 (26) | | 30 (22) | 46 (22) | | 29 (26) | 47 (19) | |
| | None | 5 (2) | 2 (5) | | 1 (1) | 6 (3) | | 2 (1) | 2 (1) | | 1 (1) | 6 (3) | |
| If self-weighed, did you have to send proof of weight | Yes | 186 (75) | 17 (81) | 0.54 | 85 (80) | 118 (72) | 0.15 | 83 (78) | 120 (74) | 0.51 | 56 (70) | 147 (78) | 0.18 |
| | No | 62 (25) | 4 (19) | | 21 (20) | 45 (28) | | 24 (22) | 42 (26) | | 24 (30) | 42 (22) | |
| **Experiences of using GLP-1 medication** | | | | | | | | | | | | |
| GLP1-RA medications are easy to use | Agree | 300 (97) | 37 (90) | **<0.01** | 119 (95) | 218 (96) | 0.93 | 128 (92) | 209 (98) | **0.02** | 105 (96) | 232 (96) | 0.72 |
| | Neutral | 9 (3) | 1 (2) | | 4 (3) | 6 (3) | | 8 (6) | 2 (1) | | 4 (4) | 6 (3) | |
| | Disagree | 2 (1) | 3 (7) | | 2 (2) | 3 (1) | | 3 (2) | 2 (2) | | 1 (1) | 4 (2) | |

| | | | | | | | | | | | | | |
|---|---|---|---|---|---|---|---|---|---|---|---|---|---|
| GLP1-RA medications are easy to access | Agree | 221 (71) | 27 (66) | 0.72 | 92 (74) | 156 (69) | 0.57 | 90 (65) | 158 (74) | **<0.001** | 74 (67) | 174 (72) | 0.58 |
| | Neutral | 44 (14) | 6 (15) | | 17 (14) | 33 (15) | | 17 (12) | 33 (16) | | 16 (15) | 34 (14) | |
| | Disagree | 46 (15) | 8 (20) | | 16 (13) | 38 (17) | | 32 (23) | 22 (10) | | 20 (18) | 34 (14) | |
| GLP-1 medications improved my overall quality of life | Agree | 271 (87) | 35 (85) | 0.48 | 106 (85) | 200 (88) | 0.9 | 117 (84) | 189 (89) | 0.40 | 84 (76) | 222 (92) | **<0.001** |
| | Neutral | 33 (11) | 6 (15) | | 15 (12) | 24 (11) | | 18 (13) | 21 (10) | | 23 (21) | 16 (7) | |
| | Disagree | 7 (2) | 0 (0) | | 4 (3) | 3 (1) | | 4 (3) | 3 (1) | | 3 (3) | 4 (2) | |
| I experienced significant side effects while using GLP1-RA medications | Agree | 86 (28) | 16 (39) | 0.09 | 38 (30) | 64 (28) | 0.84 | 38 (27) | 64 (30) | 0.81 | 33 (30) | 69 (29) | 0.67 |
| | Neutral | 54 (17) | 10 (24) | | 21 (17) | 43 (19) | | 27 (19) | 37 (17) | | 17 (16) | 47 (19) | |
| | Disagree | 171 (55) | 15 (37) | | 66 (53) | 120 (53) | | 74 (53) | 112 (53) | | 60 (55) | 126 (52) | |
| Since starting GLP1-RA medication, how has your physical activity level changed? | Increased | 194 (62) | 25 (61) | 0.97 | 73 (58) | 146 (64) | 0.49 | 84 (60) | 135 (63) | **0.053** | 45 (41) | 174 (72) | **<0.001** |
| | Stayed the same | 104 (33) | 14 (34) | | 47 (38) | 71 (31) | | 53 (38) | 65 (31) | | 61 (56) | 57 (24) | |
| | Decreased | 13 (4) | 2 (5) | | 5 (4) | 10 (4) | | 2 (1) | 13 (6) | | 4 (4) | 11 (5) | |
| Since starting GLP1-RAs, how has your mental health changed? | Improved | 177 (57) | 28 (68) | 0.20 | 73 (58) | 132 (58) | 0.93 | 78 (56) | 127 (60) | 0.29 | 52 (47) | 153 (63) | **0.02** |
| | Stayed the same | 126 (41) | 11 (27) | | 49 (39) | 88 (39) | | 59 (42) | 78 (37) | | 54 (49) | 83 (34) | |

| | | | | | | | | | | | | | |
|---|---|---|---|---|---|---|---|---|---|---|---|---|---|
| | Worsened | 8 (3) | 2 (5) | | 3 (2) | 7 (3) | | 2 (1) | 8 (4) | | 4 (4) | 6 (3) | |
| How has the quantity of food you consume been affected? | Less food | 301 (97) | 37 (90) | **<0.001** | 121 (97) | 217 (96) | 0.32 | 132 (95) | 206 (97) | 0.34 | 103 (94) | 235 (97) | 0.30 |
| | No change | 9 (3) | 1 (2) | | 3 (3) | 6 (3) | | 4 (3) | 6 (3) | | 5 (5) | 5 (2) | |
| | More food | 1 (<1) | 3 (7) | | 0 (0) | 4 (2) | | 3 (2) | 1 (1) | | 2 (2) | 2 (1) | |
| Do you feel GLP1-RA has improved the quality of your food choices? | Healthier | 244 (79) | 29 (71) | 0.47 | 100 (80) | 173 (76) | 0.71 | 110 (79) | 163 (77) | 0.56 | 79 (72) | 194 (80) | 0.22 |
| | Stayed the same | 59 (19) | 10 (24) | | 22 (18) | 47 (21) | | 24 (17) | 45 (21) | | 27 (25) | 42 (17) | |
| | Less healthy | 8 (3) | 2 (5) | | 3 (2) | 7 (3) | | 5 (4) | 5 (2) | | 4 (4) | 6 (3) | |
| **Healthcare provider support** | | | | | | | | | | | | | |
| My healthcare provider is knowledgeable about GLP-1RA medication | Agree | 228 (73) | 33 (81) | 0.58 | 94 (75) | 167 (74) | 0.81 | 100 (72) | 161 (76) | 0.42 | 77 (70) | 184 (76) | 0.49 |
| | Neutral | 76 (24) | 7 (17) | | 29 (23) | 54 (24) | | 37 (27) | 46 (22) | | 30 (27) | 53 (22) | |
| | Disagree | 7 (2) | 1 (2) | | 2 (2) | 6 (3) | | 2 (1) | 6 (3) | | 3 (3) | 5 (2) | |
| My healthcare provider provides adequate support | Agree | 191 (61) | 26 (63) | 0.93 | 76 (61) | 141 (62) | 0.96 | 87 (63) | 130 (61) | 0.92 | 64 (58) | 153 (63) | 0.14 |
| | Neutral | 92 (30) | 11 (27) | | 37 (30) | 66 (29) | | 39 (28) | 64 (30) | | 31 (28) | 72 (30) | |
| | Disagree | 28 (9) | 4 (10 | | 12 (10) | 20 (9) | | 13 (9) | 19 (9) | | 15 (14) | 17 (7) | |

| | | | | | | | | | | | | | |
|---|---|---|---|---|---|---|---|---|---|---|---|---|---|
| My healthcare provider provides adequate dietary advice during my treatment | Agree | 147 (47) | 24 (59) | 0.33 | 60 (48) | 111 (49) | 0.45 | 68 (49) | 103 (48) | 0.99 | 52 (47) | 119 (49) | 0.59 |
| | Neutral | 92 (30) | 11 (27) | | 41 (33) | 62 (27) | | 40 (29) | 63 (30) | | 30 (27) | 73 (30) | |
| | Disagree | 72 (23) | 6 (15) | | 24 (19) | 54 (24) | | 31 (22) | 47 (22) | | 28 (26) | 50 (21) | |
| My healthcare provider provides adequate physical activity guidance | Agree | 111 (36) | 16 (39) | 0.65 | 45 (36) | 82 (36) | 0.40 | 53 (38) | 74 (35) | 0.68 | 36 (33) | 91 (38) | 0.67 |
| | Neutral | 90 (29) | 9 (22) | | 40 (32) | 59 (26) | | 40 (29) | 59 (28) | | 33 (30 | 66 (27) | |
| | Disagree | 110 (35) | 16 (39) | | 40 (32) | 86 (38) | | 46 (33) | 80 (38) | | 41 (37) | 85 (35) | |
| My healthcare provider provides adequate psychological support | Agree | 83 (27) | 13 (32) | 0.79 | 37 (30) | 59 (26) | 0.35 | 40 (29) | 56 (26) | 0.79 | 30 (27) | 66 (27) | 0.21 |
| | Neutral | 103 (33) | 12 (32) | | 45 (36) | 72 (32) | | 47 (34) | 70 (33) | | 30 (27) | 87 (36) | |
| | Disagree | 124 (40) | 15 (37) | | 43 (34) | 96 (42) | | 52 (37) | 87 (41) | | 50 (46) | 89 (37) | |
| My healthcare provider regularly monitors my weight and health | Agree | 192 (62) | 24 (59) | 0.76 | 79 (63) | 137 (60) | 0.86 | 85 (61) | 131 (62) | 0.85 | 67 (61) | 149 (62) | 0.62 |
| | Neutral | 44 (14) | 5 (12) | | 17 (14) | 32 (14) | | 21 (15) | 28 (13) | | 18 (16) | 31 (13) | |
| | Disagree | 75 (24) | 12 (29) | | 29 (23) | 58 (26) | | 33 (24) | 54 (25) | | 25 (23) | 62 (26) | |
| I feel comfortable discussing my concerns with my healthcare provider | Agree | 219 (70) | 35 (85) | **0.03** | 98 (78) | 156 (69) | 0.12 | 98 (71) | 156 (73) | 0.85 | 79 (72) | 175 (72) | 0.24 |
| | Neutral | 69 (22) | 2 (5) | | 21 (17) | 50 (22) | | 30 (22) | 41 (19) | | 19 (17) | 52 (22) | |

| | | | | | | | | | | | | | |
|---|---|---|---|---|---|---|---|---|---|---|---|---|---|
| | Disagree | 23 (7) | 4 (10) | | 6 (5) | 21 (9) | | 11 (8) | 16 (8) | | 12 (11) | 15 (6) | |
| **Barriers to adherence** | | | | | | | | | | | | | |
| **Cost of medication** | Extremely challenging | 96 (31) | 11 (27) | 0.84 | 41 (33) | 66 (29) | 0.87 | 52 (37) | 55 (26) | 0.10 | 27 (25) | 80 (33) | 0.12 |
| | Moderately challenging | 115 (37) | 16 (39) | | 44 (35) | 87 (38) | | 43 (31) | 88 (41) | | 41 (37) | 90 (37) | |
| | Slightly challenging | 46 (15) | 5 (12) | | 17 (14) | 34 (15) | | 19 (14) | 32 (15) | | 15 (14) | 36 (15) | |
| | Not challenging | 54 (17) | 9 (22) | | 23 (18) | 40 (18) | | 25 (18) | 38 (18) | | 27 (25) | 36 (15) | |
| **Side effects** | Extremely challenging | 18 (6) | 2 (5) | 0.45 | 9 (7) | 11 (5) | 0.62 | 7 (5) | 13 (6) | 0.30 | 8 (7) | 12 (5) | 0.73 |
| | Moderately challenging | 52 (17) | 11 (27) | | 25 (20) | 38 (17) | | 23 (17) | 40 (19) | | 17 (16) | 46 (19) | |
| | Slightly challenging | 112 (36) | 14 (34) | | 41 (33) | 85 (37) | | 44 (32) | 82 (39) | | 39 (36) | 87 (36) | |
| | Not challenging | 129 (42 | 14 (34) | | 50 (40) | 93 (41 | | 65 (47) | 78 (37) | | 46 (42) | 97 (40) | |
| **Access to medication** | Extremely challenging | 11 (4) | 2 (5) | 0.28 | 1 (1) | 12 (5) | 0.06 | 6 (4) | 7 (3) | 0.79 | 5 (5) | 8 (3) | 0.23 |
| | Moderately challenging | 42 (14) | 10 (24) | | 17 (14) | 35 (15) | | 20 (14) | 32 (15) | | 10 (9) | 42 (17) | |
| | Slightly challenging | 80 (26) | 8 (20) | | 27 (22) | 61 (27) | | 38 (27) | 50 (24) | | 29 (26) | 59 (24) | |
| | Not challenging | 178 (57) | 21 (51) | | 80 (64) | 119 (52) | | 75 (54) | 124 (58) | | 66 (60) | 133 (55) | |
| **Social stigma** | Extremely challenging | 21 (7) | 6 (15) | 0.34 | 10 (8) | 17 (8) | 0.41 | 12 (9) | 15 (7) | 0.07 | 7 (6) | 20 (8) | **0.02** |
| | Moderately challenging | 60 (19) | 8 (20) | | 24 (19) | 44 (19) | | 24 (17) | 44 (21) | | 12 (11) | 56 (23) | |

| | | | | | | | | | | | | | |
|---|---|---|---|---|---|---|---|---|---|---|---|---|---|
| | Slightly challenging | 78 (25) | 10 (24) | | 25 (20) | 63 (28) | | 26 (19) | 62 (29) | | 27 (25) | 61 (25) | |
| | Not challenging | 152 (49) | 17 (42) | | 66 (53) | 103 (45) | | 77 (54) | 92 (43) | | 64 (58) | 105 (43) | |
| **Negative effects on social life** | Extremely challenging | 11 (4) | 2 (5) | 0.28 | 3 (2) | 10 (4) | 0.46 | 5 (4) | 8 (4) | 0.26 | 3 (3) | 10 (4) | 0.41 |
| | Moderately challenging | 37 (12) | 9 (22) | | 15 (12) | 31 (14) | | 15 (11) | 31 (15) | | 10 (9) | 36 (15) | |
| | Slightly challenging | 91 (29) | 12 (29 | | 33 (26) | 70 (31) | | 35 (25) | 68 (32) | | 35 (32) | 68 (28) | |
| | Not challenging | 172 (55) | 19 (44) | | 74 (59) | 116 (51) | | 84 (60) | 106 (50) | | 62 (56) | 128 (53) | |
| **Motivators to adherence** | | | | | | | | | | | | | |
| **Improved health outcomes** | Extremely important | 212 (68) | 25 (61) | 0.73 | 84 (67) | 153 (67) | 0.76 | 89 (64) | 148 (70) | 0.76 | 74 (67) | 163 (67) | 0.15 |
| | Moderately important | 87 (28) | 14 (34) | | 37 (30) | 64 (28) | | 44 (32) | 57 (27) | | 29 (26) | 72 (30) | |
| | Slightly important | 10 (3) | 2 (5) | | 4 (3) | 8 (4) | | 5 (4) | 7 (3) | | 7 (6) | 5 (2) | |
| | Not important | 2 (1) | 0 (0) | | 0 (0) | 2 (1) | | 1 (1) | 1 (1) | | 0 (0) | 2 (1) | |
| **Weight loss** | Extremely important | 274 (88) | 32 (78) | 0.15 | 105 (84) | 201 (89) | 0.45 | 115 (83) | 191 (90) | 0.13 | 98 (89) | 208 (86) | 0.21 |
| | Moderately important | 35 (11) | 8 (20) | | 19 (15) | 24 (11) | | 23 (17) | 20 (9) | | 10 (9) | 33 (14) | |
| | Slightly important | 2 (1) | 1 (2) | | 1 (1) | 2 (1) | | 1 (1) | 2 (1) | | 2 (2) | 1 (<1) | |
| | Not important | 0 (0) | 0 (0) | | 0 (0) | 0 (0) | | 0 (0) | 0 (0) | | 0 (0) | 0 (0) | |

| | | | | | | | | | | | | | |
|---|---|---|---|---|---|---|---|---|---|---|---|---|---|
| **Improved body image** | Extremely important | 191 (61) | 26 (63) | 0.36 | 73 (58) | 144 (63) | **0.01** | 77 (55) | 140 (66) | 0.06 | 63 (57) | 154 (64) | 0.39 |
| | Moderately important | 89 (29) | 9 (22) | | 43 (34) | 55 (24) | | 43 (31) | 55 (26) | | 33 (30) | 65 (27) | |
| | Slightly important | 25 (8) | 6 (15) | | 5 (4) | 26 (12) | | 14 (10) | 17 (8) | | 13 (12) | 18 (7) | |
| | Not important | 6 (2) | 0 (0) | | 4 (3) | 2 (1) | | 5 (4) | 1 (1) | | 1 (1) | 5 (2) | |
| **Increased self confidence** | Extremely important | 171 (55) | 19 (46) | 0.21 | 68 (54) | 122 (54) | 0.77 | 76 (55) | 114 (54) | 0.86 | 56 (51) | 134 (55) | 0.28 |
| | Moderately important | 100 (32) | 19 (46) | | 43 (34) | 76 (34) | | 47 (34) | 72 (34) | | 35 (32) | 84 (35) | |
| | Slightly important | 26 (8) | 3 (7) | | 8 (6) | 21 (9) | | 12 (9) | 17 (8) | | 13 (12) | 16 (7) | |
| | Not important | 14 (5) | 0 (0) | | 6 (5) | 8 (4) | | 4 (3) | 10 (5) | | 6 (6) | 8 (3) | |
| **Improved quality of life** | Extremely important | 209 (67) | 26 (63) | 0.84 | 83 (66) | 152 (67) | 0.90 | 97 (70) | 138 (65) | 0.09 | 70 (64) | 165 (68) | 0.44 |
| | Moderately important | 87 (28) | 12 (29) | | 36 (29) | 63 (28) | | 40 (29) | 59 (28) | | 32 (29) | 67 (28) | |
| | Slightly important | 14 (5) | 3 (7) | | 6 (5) | 11 (5) | | 2 (1) | 15 (7) | | 8 (7) | 9 (4) | |
| | Not important | 1 (<1) | 0 (0) | | 0 (0) | 1 (<1) | | 0 (0) | 1 (1) | | 0 (0) | 1 (<1) | |
| **Improved social life** | Extremely important | 47 (15) | 13 (32) | **<0.001** | 18 (14) | 42 (19) | 0.75 | 20 (14) | 40 (19) | 0.73 | 17 (16) | 43 (18) | 0.81 |
| | Moderately important | 57 (18) | 6 (15) | | 24 (19) | 39 (17) | | 27 (19) | 36 (17) | | 19 (17) | 44 (18) | |
| | Slightly important | 83 (27) | 15 (37) | | 34 (27) | 64 (28) | | 39 (28) | 59 (28) | | 29 (26) | 69 (29) | |
| | Not important | 124 (40) | 7 (17) | | 49 (39) | 82 (36) | | 53 (38) | 78 (37) | | 45 (41) | 86 (36) | |

| | | | | | | | | | | | | | |
|---|---|---|---|---|---|---|---|---|---|---|---|---|---|
| **Improved pain management** | Extremely important | 66 (21) | 16 (39) | 0.06 | 36 (29) | 46 (20) | 0.28 | 35 (25) | 47 (22) | 0.49 | 28 (26) | 54 (22) | 0.209 |
| | Moderately important | 77 (25) | 7 (17) | | 30 (24) | 54 (24) | | 33 (24) | 51 (24) | | 30 (27) | 54 (22) | |
| | Slightly important | 63 (20) | 9 (22) | | 24 (19) | 48 (21) | | 32 (23) | 40 (19) | | 25 (23) | 47 (19) | |
| | Not important | 105 (34) | 9 (22) | | 35 (28) | 79 (35) | | 39 (28) | 75 (35) | | 27 (25) | 87 (36) | |
| **Improved physical activity** | Extremely important | 76 (24) | 13 (32) | 0.20 | 27 (22) | 62 (27) | 0.21 | 42 (30) | 47 (22) | 0.24 | 16 (15) | 73 (30) | **<0.001** |
| | Moderately important | 128 (41) | 19 (46) | | 60 (48) | 87 (38) | | 55 (40) | 92 (43) | | 43 (39) | 104 (43) | |
| | Slightly important | 81 (26) | 9 (22) | | 27 (22) | 63 (28) | | 35 (25) | 55 (26) | | 38 (35) | 52 (22) | |
| | Not important | 26 (8) | 0 (0) | | 11 (9) | 15 (7) | | 7 (5) | 19 (9) | | 13 (12) | 13 (5) | |
| **Improved diet quality** | Extremely important | 97 (31) | 18 (44) | 0.40 | 36 (29) | 79 (35) | 0.28 | 49 (35) | 66 (31) | 0.38 | 29 (26) | 86 (36) | 0.38 |
| | Moderately important | 137 (44) | 14 (34) | | 60 (48) | 91 (40) | | 61 (44) | 90 (42) | | 50 (46) | 101 (42) | |
| | Slightly important | 54 (17) | 7 (17) | | 18 (14) | 43 (19) | | 23 (17) | 38 (18) | | 22 (20) | 39 (16) | |
| | Not important | 54 (17) | 2 (5) | | 11 (9) | 14 (6) | | 6 (4) | 19 (9) | | 9 (8) | 16 (7) | |
| **Reduction in other medication** | Extremely important | 38 (12) | 11 (27) | **0.02** | 16 (13) | 33 (15) | 0.769 | 21 (15) | 28 (13) | 0.07 | 16 (15) | 33 (14) | 0.56 |
| | Moderately important | 48 (15) | 10 (24) | | 24 (19) | 34 (15) | | 30 (22) | 28 (13) | | 22 (20) | 36 (15) | |
| | Slightly important | 43 (14) | 5 (12) | | 17 (14) | 31 (14) | | 13 (9) | 36 (16) | | 16 (15) | 32 (13) | |
| | Not important | 182 (59) | 15 (37) | | 68 (54) | 129 (57) | | 75 (54) | 122 (57) | | 56 (51) | 141 (58) | |

**Supplementary material 7. Subgroup analysis of former GLP-1 user experiences, barriers and motivators to adherence and discontinuation by age, gender and BMI.**

| Question | Response Options | Age | | | Gender | | | BMI at time of survey | | | BMI at start of treatment | | |
|---|---|---|---|---|---|---|---|---|---|---|---|---|---|
| | | **Youn-ger** | **Older** | ***p*** | **Male** | **Female** | ***p*** | **≤ 30** | **>30** | ***p*** | **25-39.9** | **40+** | ***p*** |
| **Medication use, access and provision** | | | | | | | | | | | | | |
| Type of drug | Semaglutide | 67 (45) | 66 (54) | 0.26 | 48 (57) | 84 (45) | 0.06 | 80 (47) | 53 (53) | 0.78 | 103 (50) | 30 (46) | 0.55 |
| | Tirzepatide | 74 (50) | 51 (42) | | 30 (36) | 94 (51) | | 82 (48) | 43 (43) | | 93 (45) | 1 (2) | |
| | Liraglutide | 3 (2) | 3 (2) | | 3 (4) | 5 (3) | | 6 (4) | 2 (2) | | 7 (3) | 2 (3) | |
| | Exenatide | 3 (2) | 3 (2) | | 1 (1) | 3 (2) | | 2 (1) | 2 (2) | | 2 (1) | 0 (0) | |
| Duration of use | 0-6 months | 100 (67) | 80 (65) | 0.70 | 54 (64) | 125 (67) | 0.43 | 115 (67) | 65 (64) | 0.60 | 136 (66) | 44 (68) | 0.52 |
| | 7-12 months | 34 (23) | 33 (27) | | 20 (24) | 46 (25) | | 41 (21) | 26 (26) | | 54 (26) | 13 (20) | |
| | 1-2 years | 14 (9) | 10 (8) | | 9 (11) | 15 (8) | | 15 (9) | 9 (9) | | 16 (8) | 8 (12) | |
| | 2 + years | 1 (1) | 0 (0) | | 1 (1) | 0 (0) | | 0 (0) | 1 (1) | | 1 (1) | 0 (0) | |
| Access to medication | Privately online | 95 (64) | 63 (51) | **<0.001** | 30 (36) | 127 (68) | **<0.001** | 93 (54) | 65 (64) | 0.07 | 124 (60) | 34 (52) | **0.02** |
| | Privately face to face | 16 (11) | 8 (7) | | 8 (10) | 16 (9) | | 18 (11) | 6 (6) | | 22 (11) | 2 (3) | |
| | NHS provider online | 13 (9) | 7 (6) | | 10 (12) | 10 (5) | | 17 (10) | 3 (3) | | 16 (8) | 4 (6) | |
| | NHS provider face to face | 20 (13) | 20 (13) | | 34 (41) | 28 (15) | | 37 (22) | 26 (26) | | 39 (19) | 24 (27) | |

| | | | | | | | | | | | | | |
|---|---|---|---|---|---|---|---|---|---|---|---|---|---|
| | Other | 5 (3) | 5 (3) | | 2 (2) | 5 (3) | | 6 (4) | 1 (1) | | 6 (3) | 1 (2) | |
| Which healthcare provider manages your treatment | Private healthcare | 74 (50) | 45 (37) | 0.06 | 26 (31) | 26 (14) | **<0.001** | 74 (43) | 45 (45) | 0.86 | 93 (45) | 26 (40) | 0.15 |
| | GP | 24 (16) | 37 (30) | | 34 (41) | 26 (14) | | 39 (23) | 22 (22) | | 42 (20) | 19 (29) | |
| | Specialist | 13 (9) | 11 (9) | | 12 (14) | 12 (7) | | 16 (9) | 8 (8) | | 15 (7) | 9 (14) | |
| | None | 36 (24) | 27 (22) | | 12 (14) | 50 (27) | | 40 (23) | 23 (23) | | 53 (26) | 10 (15) | |
| | Other | 2 (1) | 3 (2) | | 0 (0) | 5 (3) | | 2 (1) | 3 (3) | | 4 (2) | 1 (2) | |
| Method of confirming weight | Self-weighed | 103 (69) | 64 (52) | **0.02** | 33 (39) | 133 (72) | **<0.001** | 104 (61) | 36 (36) | 0.88 | 136 (66) | 31 (48) | **0.03** |
| | Weighed by professional | 43 (29) | 55 (45) | | 48 (57) | 49 (26) | | 62 (36) | 63 (62) | | 67 (32) | 31 (48) | |
| | None | 3 (2) | 4 (3) | | 3 (4) | 4 (2) | | 5 (3) | 2 (2) | | 4 (2) | 3 (5) | |
| If self-weighed, did you have to send proof of weight | Yes | 66 (64) | 41 (64) | 0.99 | 19 (58) | 87 (65) | 0.40 | 69 (63) | 35 (34) | 0.43 | 88 (65) | 19 (61) | 0.72 |
| | No | 37 (36) | 23 (36) | | 14 (42) | 46 (35) | | 38 (60) | 25 (40) | | 48 (35) | 12 (39) | |
| **Experiences of using GLP-1 medication** | | | | | | | | | | | | | |
| GLP1-RA medications are easy to use | Agree | 138 (93) | 115 (94) | 0.94 | 75 (89) | 176 (95) | 0.13 | 156 (91) | 97 (96) | 0.19 | 192 (93) | 61 (94) | 0.95 |
| | Neutral | 6 (4) | 4 (3) | | 6 (7) | 4 (2) | | 9 (5) | 1 (1) | | 8 (4) | 2 (3) | |
| | Disagree | 5 (3) | 4 (3) | | 3 (4) | 6 (3) | | 6 (4) | 3 (3) | | 7 (3) | 2 (3) | |

| | | | | | | | | | | | | | |
|---|---|---|---|---|---|---|---|---|---|---|---|---|---|
| GLP1-RA medications are easy to access | Agree | 113 (76) | 88 (72) | 0.41 | 63 (75) | 138 (74) | 0.99 | 119 (70) | 82 (81) | **0.02** | 152 (73) | 49 (75) | 0.07 |
| | Neutral | 16 (11) | 20 (16) | | 11 (13) | 25 (13) | | 30 (18) | 6 (6) | | 32 (16) | 4 (6) | |
| | Disagree | 20 (13) | 15 (12) | | 10 (12) | 23 (12) | | 22 (13) | 13 (13) | | 23 (11) | 12 (19) | |
| GLP-1 medications improved my overall quality of life | Agree | 89 (60) | 68 (55) | 0.73 | 59 (70) | 97 (52) | **0.02** | 114 (67) | 43 (43) | **<0.001** | 126 (61) | 31 (48) | 0.11 |
| | Neutral | 31 (21) | 27 (22) | | 13 (16) | 44 (24) | | 35 (21) | 23 (23) | | 43 (21) | 15 (23) | |
| | Disagree | 29 (20) | 28 (23 | | 12 (14) | 45 (24) | | 22 (13) | 35 (35) | | 38 (18) | 19 (29) | |
| I experienced significant side effects while using GLP1-RA medications | Agree | 61 (41) | 42 (34) | 0.47 | 20 (24) | 81 (44) | **<0.001** | 56 (33) | 47 (47) | **0.05** | 81 (39) | 22 (34) | 0.69 |
| | Neutral | 30 (20) | 25 (20) | | 19 (23) | 36 (19) | | 40 (23) | 15 (15) | | 40 (19) | 15 (23) | |
| | Disagree | 58 (39) | 56 (46) | | 45 (54) | 69 (37) | | 75 (44) | 39 (39) | | 86 (42) | 28 (43) | |
| Since starting GLP1-RA medication, how has your physical activity level changed? | Increased | 67 (45) | 56 (46) | 0.93 | 44 (52) | 78 (42) | 0.15 | 89 (52) | 34 (34) | **0.01** | 93 (45) | 30 (46) | 0.82 |
| | Stayed the same | 64 (43) | 54 (44) | | 29 (35) | 88 (47) | | 64 (37) | 54 (54) | | 89 (43) | 29 (45) | |
| | Decreased | 18 (12) | 13 (11) | | 11 (13) | 20 (11) | | 18 (11) | 13 (13) | | 25 (12) | 6 (9) | |
| Since starting GLP1-RAs, how has your mental health changed? | Improved | 63 (42) | 47 (38) | 0.73 | 37 (44) | 72 (39) | 0.26 | 77 (45) | 33 (33) | 0.11 | 86 (42) | 24 (37) | 0.55 |
| | Stayed the same | 68 (46) | 62 (50) | | 41 (49) | 88 (47) | | 77 (45) | 53 (53) | | 99 (48) | 31 (48) | |

| | | | | | | | | | | | | | |
|---|---|---|---|---|---|---|---|---|---|---|---|---|---|
| | Worsened | 18 (12) | 14 (11) | | 6 (7) | 26 (14) | | 17 (10) | 15 (15) | | 22 (11) | 10 (15) | |
| How has the quantity of food you consume been affected? | Less food | 139 (93) | 112 (91) | 0.23 | 73 (87) | 176 (95) | **0.03** | 161 (94) | 90 (89) | 0.09 | 191 (92) | 60 (92) | 0.71 |
| | No change | 8 (5) | 11 (9) | | 9 (11) | 10 (5) | | 8 (5) | 11 (11) | | 14 (7) | 5 (8) | |
| | More food | 2 (1) | 0 (0) | | 2 (2) | 0 (0) | | 2 (1) | 0 (0) | | 2 (1) | 0 (0) | |
| Do you feel GLP1-RA has improved the quality of your food choices? | Healthier | 110 (74) | 76 (62) | 0.06 | 63 (75) | 122 (66) | 0.28 | 128 (75) | 58 (57) | **<0.001** | 141 (68) | 45 (69) | 0.57 |
| | Stayed the same | 33 (22) | 43 (35) | | 18 (21) | 57 (31) | | 36 (21) | 40 (40) | | 57 (28) | 19 (29) | |
| | Less healthy | 6 (4) | 4 (3) | | 3 (4) | 7 (4) | | 7 (4) | 3 (3) | | 9 (4) | 1 (2) | |
| **Healthcare provider support** | | | | | | | | | | | | | |
| My healthcare provider is knowledgeable about GLP-1RA medication | Agree | 93 (62) | 89 (72) | 0.22 | 65 (77) | 116 (62) | **0.052** | 122 (71) | 60 (59) | 0.13 | 139 (67) | 43 (66) | 0.81 |
| | Neutral | 41 (28) | 25 (20) | | 14 (17) | 51 (27) | | 36 (21) | 30 (30) | | 51 (25) | 15 (23) | |
| | Disagree | 15 (10) | 9 (7) | | 5 (6) | 19 (10 | | 13 (8) | 11 (11) | | 17 (8) | 7 (11) | |
| My healthcare provider provides adequate support | Agree | 78 (52) | 70 (57) | 0.75 | 62 (74) | 85 (46) | **<0.001** | 106 (62) | 42 (42) | **<0.001** | 117 (57) | 31 (48) | 0.41 |
| | Neutral | 46 (31) | 34 (28) | | 16 (19) | 63 (34) | | 45 (26) | 35 (35) | | 57 (28) | 23 (35) | |
| | Disagree | 25 (17) | 19 (15) | | 6 (7) | 38 (20) | | 20 (12) | 24 (24) | | 33 (16) | 11 (17) | |

| | | | | | | | | | | | | | |
|---|---|---|---|---|---|---|---|---|---|---|---|---|---|
| My healthcare provider provides adequate dietary advice during my treatment | Agree | 71 (48) | 61 (50) | 0.86 | 58 (69) | 74 (40) | **<0.001** | 95 (56) | 37 (37) | **<0.001** | 107 (52) | 25 (39) | 0.09 |
| | Neutral | 42 (28) | 31 (25) | | 15 (18) | 56 (30) | | 47 (28) | 26 (26) | | 55 (27) | 18 (28) | |
| | Disagree | 36 (24) | 31 (25) | | 11 (13) | 56 (30) | | 29 (17) | 38 (38) | | 45 (22) | 22 (34) | |
| My healthcare provider provides adequate physical activity guidance | Agree | 56 (38) | 45 (37) | 0.90 | 50 (60) | 51 (27) | **<0.001** | 73 (43) | 28 (28) | **<0.001** | 79 (38) | 22 (34) | 0.11 |
| | Neutral | 46 (31) | 36 (29) | | 19 (23) | 61 (33) | | 59 (35) | 23 (23) | | 67 (32) | 15 (23) | |
| | Disagree | 47 (32) | 42 (34) | | 15 (18) | 74 (40) | | 39 (23) | 50 (50) | | 61 (30) | 28 (43) | |
| My healthcare provider provides adequate psychological support | Agree | 47 (32) | 30 (24) | 0.37 | 42 (50) | 35 (19) | **<0.001** | 59 (35) | 18 (18) | **<0.001** | 63 (30) | 14 (22) | 0.12 |
| | Neutral | 41 (28) | 41 (33) | | 21 (25) | 60 (32) | | 57 (33) | 25 (25) | | 65 (31) | 17 (26) | |
| | Disagree | 61 (41) | 52 (42) | | 21 (25) | 91 (49) | | 55 (32) | 58 (57) | | 79 (38) | 34 (52) | |
| My healthcare provider regularly monitors my weight and health | Agree | 76 (51) | 65 (53) | 0.96 | 55 (66) | 85 (46) | **<0.001** | 100 (59) | 41 (41) | **0.02** | 108 (52) | 33 (51) | 0.76 |
| | Neutral | 29 (20) | 23 (19) | | 17 (20) | 35 (19) | | 29 (17) | 23 (23) | | 41 (20) | 11 (17) | |
| | Disagree | 44 (30) | 35 (29) | | 12 (14) | 66 (36) | | 42 (25) | 37 (37) | | 58 (28) | 21 (32) | |
| I feel comfortable discussing my concerns with my healthcare provider | Agree | 96 (64) | 85 (69) | 0.60 | 70 (83) | 111 (60) | <0.001 | 119 (70) | 62 (61) | 0.25 | 135 (65) | 46 (71) | 0.71 |
| | Neutral | 38 (26) | 25 (20) | | 6 (7) | 55 (30) | | 38 (22) | 25 (25) | | 50 (24) | 13 (20) | |

| | | | | | | | | | | | | | |
|---|---|---|---|---|---|---|---|---|---|---|---|---|---|
| | Disagree | 15 (10) | 13 (11) | | 8 (10) | 20 (11) | | 14 (8) | 14 (14) | | 22 (11) | 6 (9) | |
| **Barriers to adherence** | | | | | | | | | | | | | |
| **Cost of medication** | Extremely challenging | 44 (30) | 45 (37) | 0.54 | 11 (13) | 77 (41) | **<0.001** | 49 (29) | 40 (40) | **0.04** | 65 (31) | 24 (37) | 0.13 |
| | Moderately challenging | 55 (37) | 40 (33) | | 36 (43) | 59 (32) | | 63 (37) | 32 (32) | | 75 (36) | 20 (31) | |
| | Slightly challenging | 24 (16) | 15 (12) | | 16 (19) | 23 (12) | | 31 (18) | 8 (8) | | 34 (16) | 5 (8) | |
| | Not challenging | 26 (17) | 23 (19) | | 21 (25) | 27 (15) | | 28 (16) | 21 (21) | | 33 (16) | 16 (25) | |
| **Side effects** | Extremely challenging | 37 (25) | 22 (18) | 0.40 | 10 (12) | 49 (26) | **0.051** | 27 (16) | 32 (32) | **0.01** | 42 (20) | 17 (26) | 0.52 |
| | Moderately challenging | 28 (19) | 23 (19) | | 17 (20) | 32 (17) | | 35 (21) | 16 (16) | | 39 (19) | 12 (19) | |
| | Slightly challenging | 45 (30) | 36 (29) | | 26 (31) | 55 (30) | | 58 (34) | 23 (23) | | 66 (32) | 15 (23) | |
| | Not challenging | 39 (26) | 42 (34) | | 31 (37) | 50 (27) | | 51 (30) | 30 (30) | | 60 (29) | 21 (32) | |
| **Access to medication** | Extremely challenging | 9 (6) | 13 (11) | 0.52 | 4 (5) | 17 (9) | 0.15 | 11 (6) | 11 (11) | **0.03** | 11 (5) | 11 (17) | **<0.001** |
| | Moderately challenging | 22 (15) | 19 (15) | | 11 (13) | 29 (16) | | 31 (18) | 10 (10 | | 33 (16) | 8 (12) | |
| | Slightly challenging | 35 (24) | 24 (20) | | 25 (30) | 34 (18) | | 43 (25) | 16 (16) | | 52 (25) | 7 (11) | |
| | Not challenging | 83 (56) | 67 (55) | | 44 (52) | 106 (57) | | 86 (50) | 64 (63) | | 111 (54) | 39 (60) | |
| **Social stigma** | Extremely challenging | 8 (5) | 4 (3) | 0.74 | 2 (2) | 10 (5) | 0.44 | 8 (5) | 4 (4) | 0.62 | 8 (4) | 4 (6) | 0.76 |
| | Moderately challenging | 17 (11) | 11 (9) | | 11 (13) | 17 (9) | | 19 (11) | 9 (9) | | 23 (11) | 5 (8) | |

| | | | | | | | | | | | | | |
|---|---|---|---|---|---|---|---|---|---|---|---|---|---|
| | Slightly challenging | 32 (22) | 28 (23) | | 21 (25) | 39 (21) | | 41 (24) | 19 (19) | | 46 (22) | 14 (22) | |
| | Not challenging | 92 (62) | 80 (65) | | 50 (60) | 120 (65) | | 103 (60) | 69 (68) | | 130 (63) | 42 (65) | |
| **Negative effects on social life** | Extremely challenging | 9 (6) | 1 (1) | 0.09 | 1 (1) | 9 (5) | 0.23 | 5 (3) | 5 (5) | 0.59 | 9 (4) | 1 (2) | 0.77 |
| | Moderately challenging | 18 (12) | 18 (15) | | 15 (18) | 21 (11) | | 25 (15) | 11 (11) | | 27 (13) | 9 (14) | |
| | Slightly challenging | 43 (29) | 30 (24) | | 20 (24) | 52 (28) | | 48 (28) | 25 (25) | | 55 (27) | 18 (28) | |
| | Not challenging | 79 (53) | 74 (60) | | 48 (57) | 104 (56) | | 93 (54) | 60 (59) | | 116 (56) | 37 (57) | |
| **Motivators to adherence** | | | | | | | | | | | | | |
| **Improved health outcomes** | Extremely important | 73 (49) | 63 (51) | 0.15 | 39 (46) | 97 (52) | 0.75 | 79 (46) | 57 (56) | 0.15 | 98 (47) | 38 (59) | 0.24 |
| | Moderately important | 52 (35) | 44 (36) | | 30 (36) | 64 (34) | | 61 (36) | 35 (35) | | 74 (36) | 22 (34) | |
| | Slightly important | 21 (14) | 9 (7) | | 11 (13) | 19 (10) | | 24 (14) | 6 (6) | | 26 (13) | 4 (6) | |
| | Not important | 3 (2) | 7 (6) | | 4 (5) | 6 (3) | | 7 (4) | 3 (3) | | 9 (4) | 1 (2) | |
| **Weight loss** | Extremely important | 121 (81) | 96 (78) | 0.21 | 55 (66) | 160 (86) | **<0.001** | 133 (78) | 84 (83) | 0.69 | 164 (79) | 53 (82) | 0.80 |
| | Moderately important | 26 (17) | 20 (16) | | 23 (17) | 23 (12) | | 32 (19) | 14 (14) | | 35 (17) | 11 (17) | |
| | Slightly important | 2 (1) | 5 (4) | | 4 (5) | 3 (2) | | 5 (3) | 2 (2) | | 6 (3) | 1 (2) | |
| | Not important | 0 (0) | 2 (2) | | 2 (2) | 0 (0) | | 1 (1) | 1 (1) | | 2 (1) | 0 (0) | |
| **Improved body image** | Extremely important | 84 (56) | 60 (49) | 0.14 | 29 (35) | 114 (61) | **<0.001** | 95 (56) | 49 (49) | 0.58 | 113 (55) | 31 (48) | 0.52 |

| | | | | | | | | | | | | | |
|---|---|---|---|---|---|---|---|---|---|---|---|---|---|
| | Moderately important | 49 (33) | 39 (32) | | 31 (37) | 56 (30) | | 54 (32) | 34 (34) | | 67 (32) | 21 (32) | |
| | Slightly important | 12 (8) | 14 (11) | | 15 (18) | 11 (6) | | 15 (9) | 11 (11) | | 17 (8) | 9 (14) | |
| | Not important | 4 (3) | 10 (8) | | 9 (11) | 5 (3) | | 7 (4) | 7 (7) | | 10 (5) | 4 (6) | |
| **Increased self confidence** | Extremely important | 77 (52) | 49 (40) | 0.24 | 26 (31) | 100 (54) | **<0.001** | 80 (47) | 45 (46) | 0.45 | 100 (48) | 26 (40) | 0.43 |
| | Moderately important | 51 (34) | 49 (40) | | 36 (43) | 62 (33) | | 35 (35) | 35 (35) | | 75 (36) | 25 (39) | |
| | Slightly important | 15 (10) | 17 (14) | | 14 (17) | 18 (10) | | 12 (12) | 12 (12) | | 21 (10) | 11 (17) | |
| | Not important | 6 (4) | 8 (7) | | 8 (10) | 6 (3) | | 8 (8) | 8 (8) | | 11 (5) | 3 (5) | |
| **Improved quality of life** | Extremely important | 76 (51) | 58 (47) | 0.39 | 31 (37) | 102 (55) | **0.04** | 79 (46) | 55 (55) | 0.14 | 97 (47) | 37 (57) | 0.42 |
| | Moderately important | 45 (30) | 45 (37) | | 33 (39) | 56 (30) | | 55 (32) | 35 (35) | | 70 (34) | 20 (31) | |
| | Slightly important | 24 (16) | 14 (11) | | 15 (18) | 23 (12) | | 30 (18) | 8 (8) | | 31 (15) | 7 (11) | |
| | Not important | 4 (3) | 6 (5) | | 5 (6) | 5 (3) | | 7 (4) | 3 (3) | | 9 (4) | 1 (2) | |
| **Improved social life** | Extremely important | 32 (22) | 16 (13) | 0.31 | 14 (17) | 34 (18) | 0.92 | 31 (18) | 17 (17) | 0.88 | 37 (18) | 11 (17) | 0.87 |
| | Moderately important | 27 (18) | 29 (23) | | 19 (23) | 36 (19) | | 32 (19) | 23 (23) | | 43 (21) | 12 (19) | |
| | Slightly important | 37 (25) | 32 (26) | | 20 (24) | 48 (26) | | 44 (26) | 25 (25) | | 50 (24) | 19 (29) | |
| | Not important | 53 (36) | 47 (38) | | 31 (37) | 68 (37) | | 64 (37) | 36 (36) | | 77 (37) | 23 (35) | |
| **Improved pain management** | Extremely important | 20 (13) | 23 (19) | 0.39 | 12 (14) | 31 (17) | 0.48 | 20 (12) | 23 (23) | **0.03** | 29 (14) | 14 (22) | **<0.001** |

| | | | | | | | | | | | | | |
|---|---|---|---|---|---|---|---|---|---|---|---|---|---|
| | Moderately important | 26 (17) | 26 (21) | | 17 (20) | 34 (18) | | 29 (17) | 23 (23) | | 32 (16) | 20 (31) | |
| | Slightly important | 30 (20) | 18 (15) | | 19 (23) | 29 (16) | | 34 (20) | 14 (14) | | 34 (16) | 14 (22) | |
| | Not important | 73 (49) | 56 (46) | | 36 (43) | 92 (50) | | 88 (52) | 41 (41) | | 112 (54) | 17 (26) | |
| **Improved physical activity** | Extremely important | 33 (22) | 28 (23) | 0.61 | 21 (25) | 39 (21) | 0.59 | 39 (23) | 22 (22) | 0.20 | 50 (24) | 11 (17) | 0.13 |
| | Moderately important | 52 (35) | 51 (42) | | 34 (41) | 69 (37) | | 60 (35) | 43 (43) | | 71 (34) | 32 (49) | |
| | Slightly important | 36 (24) | 23 (19) | | 14 (17) | 44 (24) | | 25 (21) | 24 (24) | | 45 (22) | 14 (22) | |
| | Not important | 28 (19) | 21 (17) | | 15 (18) | 34 (18) | | 37 (22) | 12 (12) | | 41 (20) | 8 (12) | |
| **Improved diet quality** | Extremely important | 50 (34) | 33 (27) | 0.64 | 24 (29) | 59 (32) | 0.57 | 55 (32) | 28 (28) | 0.10 | 62 (30) | 21 (32) | 0.68 |
| | Moderately important | 50 (34) | 44 (36) | | 28 (33) | 65 (35) | | 50 (29) | 44 (44) | | 69 (33) | 25 (39) | |
| | Slightly important | 32 (22) | 28 (23) | | 23 (27) | 37 (20) | | 43 (25) | 17 (17) | | 47 (23) | 13 (20) | |
| | Not important | 17 (11) | 18 (15) | | 9 (11) | 25 (13) | | 23 (14) | 12 (12) | | 29 (14) | 6 (9) | |
| **Reduction in other medication** | Extremely important | 8 (5) | 12 (10) | 0.17 | 10 (12) | 10 (5) | **0.02** | 10 (6) | 10 (10) | 0.40 | 13 (6) | 7 (11) | 0.53 |
| | Moderately important | 19 (13) | 24 (20) | | 19 (23) | 23 (12) | | 28 (16) | 15 (15) | | 31 (15) | 12 (19) | |
| | Slightly important | 24 (16) | 15 (12) | | 13 (16) | 26 (14) | | 28 (16) | 11 (11) | | 31 (15) | 8 (12) | |
| | Not important | 98 (66) | 72 (59) | | 42 (50) | 127 (68) | | 105 (61) | 65 (64) | | 132 (64) | 38 (59) | |
| **Discontinuation and post-treatment impact** | | | | | | | | | | | | | |

| | | | | | | | | | | | | | |
|---|---|---|---|---|---|---|---|---|---|---|---|---|---|
| **Reason for stopping GLP-1 medication** | Cost of treatment | 48 (32) | 35 (29) | 0.61 | 12 (14) | 70 (38) | **<0.001** | 52 (30) | 31 (31) | **<0.001** | 69 (33) | 14 (22) | 0.26 |
| | Planned end of treatment | 37 (25) | 30 (24) | | 42 (50) | 25 (13) | | 57 (33) | 10 (1) | | 54 (26) | 13 (20) | |
| | Side effects | 38 (26) | 31 (25) | | 10 (12) | 58 (31) | | 31 (18) | 38 (38) | | 48 (23) | 21 (32) | |
| | Lack of motivation/ben-efit | 3 (2) | 9 (7) | | 5 (6) | 7 (4) | | 6 (4) | 6 (6) | | 7 (3) | 5 (8) | |
| | Accessibility issues | 4 (3) | 3 (2) | | 3 (4) | 4 (2) | | 4 (2) | 3 (3) | | 5 (2) | 2 (3) | |
| | Stigma | 2 (1) | 1 (1) | | 2 (2) | 1 (0) | | 3 (2) | 0 (0) | | 3 (1) | 0 (0) | |
| | Changes to social life | 1 (1) | 2 (2) | | 2 (2) | 2 (2) | | 2 (1) | 1 (1) | | 2 (1) | 1 (2) | |
| | Other | 16 (11) | 12 (10) | | 8 (10) | 20 (11) | | 16 (9) | 12 (12) | | 19 (9) | 9 (14) | |
| **Barriers to restarting GLP-1 medication** | Cost of treatment | 35 (24) | 24 (20) | 0.49 | 19 (23) | 70 (38) | **<0.001** | 52 (30) | 38 (38) | **<0.001** | 69 (33) | 21 (32) | 0.54 |
| | Planned end of treatment | 40 (27) | 29 (24) | | 35 (42) | 34 (18) | | 60 (35) | 9 (9) | | 55 (27) | 14 (22) | |

| | | | | | | | | | | | | | |
|---|---|---|---|---|---|---|---|---|---|---|---|---|---|
| | Side effects | 35 (24) | 24 (20) | | 10 (12) | 49 (26) | | 27 (16) | 32 (32) | | 44 (21) | 15 (23) | |
| | Lack of motivation/ben-efit | 12 (8) | 14 (11) | | 15 (18) | 11 (6) | | 17 (10) | 9 (9) | | 21 (10) | 5 (8) | |
| | Accessibility issues | 5 (3) | 6 (5) | | 1 (1) | 9 (5) | | 6 (4) | 5 (5) | | 6 (3) | 5 (8) | |
| | Stigma | 0 (0) | 0 (0) | | 0 (0) | 0 (0) | | 0 (0) | 0 (0) | | 0 (0) | 0 (0) | |
| | Changes to social life | 0 (0) | 0 (0) | | 2 (2) | 0 (0) | | 2 (1) | 0 (0) | | 2 (1) | 0 (0) | |
| | Other | 10 (7) | 5 (4) | | 2 (2) | 13 (7) | | 7 (4) | 8 (8) | | 10 (5) | 5 (8) | |
| **What happened to your weight after you stopped taking GLP-1 medication?** | Gained weight | 75 (50) | 48 (39) | 0.07 | 24 (29) | 97 (52) | **<0.001** | 72 (42) | 51 (51) | 0.20 | 94 (45) | 29 (45) | 0.98 |
| | Maintained | 51 (34) | 59 (48) | | 42 (50) | 68 (37) | | 70 (41) | 40 (40) | | 83 (40) | 27 (42) | |
| | Lost weight | 23 (15) | 16 (13) | | 18 (21) | 21 (11) | | 29 (17) | 10 (10) | | 30 (15) | 9 (14) | |
| **What happened to your other health conditions after** | Improvement | 18 (23) | 21 (26) | 0.86 | 22 (36) | 17 (18) | **0.01** | 24 (25) | 15 (24) | 0.47 | 25 (22) | 14 (30) | 0.24 |

| | | | | | | | | | | | | | |
|---|---|---|---|---|---|---|---|---|---|---|---|---|---|
| **you stopped taking GLP-1 medication?** | No change | 54 (68) | 52 (64) | | 38 (61) | 67 (70) | | 67 (68) | 39 (63) | | 80 (70) | 26 (57) | |
| | Worsening | 7 (9) | 8 (10) | | 2 (3) | 12 (13) | | 7 (7) | 8 (13) | | 9 (8) | 6 (13) | |
| **What happened to your physical activity after you stopped taking GLP-1 medication?** | Increased | 48 (32) | 43 (35) | 0.16 | 39 (46) | 52 (28) | **0.01** | 65 (38) | 26 (26) | 0.11 | 71 (34) | 20 (31) | 0.70 |
| | No change | 84 (56) | 74 (60) | | 39 (46) | 117 (63) | | 93 (54) | 65 (64) | | 120 (58) | 38 (59) | |
| | Decreased | 17 (11) | 6 (5) | | 6 (7) | 17 (9) | | 13 (8) | 10 (10) | | 16 (8) | 7 (11) | |
| **What happened to your mental health after you stopped taking GLP-1 medication?** | Improvement | 30 (20) | 29 (24) | 0.76 | 25 (30) | 34 (18) | **0.03** | 41 (24) | 18 (18) | 0.15 | 45 (22) | 14 (22) | 0.68 |
| | No change | 93 (62) | 75 (61) | | 51 (61) | 116 (62) | | 107 (63) | 61 (60) | | 130 (63) | 38 (59) | |
| | Worsening | 26 (17) | 19 (15) | | 8 (10) | 36 (19) | | 23 (14) | 22 (22) | | 32 (16) | 13 (20) | |
| **What happened to the quantity of food you consumed when you stopped taking GLP-1 medication?** | Increased | 30 (20) | 26 (21) | 0.53 | 32 (38) | 24 (13) | <0.001 | 48 (28) | 8 (8) | **<0.001** | 46 (22) | 10 (15) | 0.39 |
| | No change | 31 (21) | 32 (26) | | 24 (29) | 39 (21) | | 34 (20) | 29 (29) | | 45 (22) | 18 (28) | |

| | | | | | | | | | | | | | |
|---|---|---|---|---|---|---|---|---|---|---|---|---|---|
| | Decreased | 88 (59) | 65 (53) | | 28 (33) | 123 (66) | | 89 (52) | 64 (64) | | 116 (56) | 37 (57) | |
| **What happened to the quality of food you consumed when you stopped taking GLP-1 medication?** | Increased | 53 (36) | 42 (34) | | 39 (46) | 56 (30) | | 71 (42) | 24 (24) | | 72 (35) | 23 (25) | |
| | No change | 73 (49) | 67 (55) | 0.54 | 36 (43) | 103 (55) | 0.03 | 80 (47) | 60 (59) | **0.01** | 112 (54) | 28 (43) | 0.08 |
| | Decreased | 23 (15) | 14 (11) | | 9 (11) | 27 (15) | | 20 (12) | 17 (17) | | 23 (11) | 14 (22) | |

**Supplementary table 8. Subgroup analysis of former GLP-1 user experiences, barriers and motivators to adherence and discontinuation by ethnicity, education, social status and physical activity.**

| Question | Response Options | Ethnicity | | | Education | | | Social status | | | Physical activity | | |
|---|---|---|---|---|---|---|---|---|---|---|---|---|---|
| | | White | Other | *p* | Lower | Higher | *p* | Lower | Higher | *p* | No | Yes | *p* |
| **Medication use, access and provision** | | | | | | | | | | | | | |
| Type of drug | Semaglutide | 130 (49) | 16 (40) | 0.55 | 46 (47) | 87 (50) | 0.42 | 48 (50) | 85 (49) | 0.40 | 36 (47) | 97 (50) | 0.90 |
| | Tirzepatide | 119 (45) | 23 (58) | | 45 (46) | 80 (46) | | 41 (42) | 84 (48) | | 37 (49) | 88 (45) | |
| | Liraglutide | 8 (3) | 1 (3) | | 5 (5) | 3 (2) | | 4 (4) | 4 (2) | | 2 (3) | 6 (3) | |
| | Exenatide | 4 (2) | 0 (0) | | 1 (1) | 3 (2) | | 3 (3) | 1 (1) | | 1 (1) | 3 (2) | |
| Duration of use | 0-6 months | 149 (65) | 30 (75) | 0.60 | 62 (64) | 118 (67) | **0.03** | 65 (67) | 115 (66) | 0.76 | 46 (61) | 134 (68) | 0.52 |
| | 7-12 months | 58 (25) | 8 (20) | | 20 (21) | 47 (27) | | 22 (23) | 45 (26) | | 23 (30) | 44 (22) | |
| | 1-2 years | 22 (10) | 2 (5) | | 15 (16) | 9 (5) | | 10 (10) | 14 (8) | | 7 (9) | 17 (9) | |
| | 2 + years | 1 (<1) | 0 (0) | | 0 (0) | 1 (1) | | 0 (0) | 0 (0) | | 0 (0) | 1 (1) | |
| Access to medication | Privately online | 142 (62) | 15 (38) | **<0.001** | 53 (55) | 105 (60) | 0.10 | 59 (61) | 99 (57) | **0.01** | 48 (63) | 110 (56) | 0.10 |
| | Privately face to face | 14 (6) | 9 (23) | | 5 (5) | 19 (11) | | 2 (2) | 22 (13) | | 5 (7) | 19 (10) | |
| | NHS provider online | 13 (6) | 7 (18) | | 9 (9) | 11 (6) | | 4 (4) | 16 (9) | | 9 (12) | 11 (6) | |
| | NHS provider face to face | 55 (24) | 8 (20) | | 25 (26) | 38 (22) | | 29 (30) | 34 (19) | | 14 (18) | 49 (25) | |

| | | | | | | | | | | | | | |
|---|---|---|---|---|---|---|---|---|---|---|---|---|---|
| | Other | 6 (3) | 1 (3) | | 5 (5) | 2 (1) | | 3 (3) | 4 (2) | | 0 (0) | 7 (4) | |
| Which healthcare provider manages your treatment | Private healthcare | 101 (44) | 18 (45) | 0.31 | 35 (36) | 84 (48) | 0.16 | 36 (37) | 63 (47) | **0.02** | 31 (41) | 88 (45) | 0.41 |
| | GP | 51 (22) | 10 (25) | | 26 (27) | 35 (20) | | 25 (26) | 36 (21) | | 19 (25) | 42 (21) | |
| | Specialist | 17 (7) | 6 (15) | | 6 (6) | 18 (10) | | 9 (9) | 15 (9) | | 5 (7) | 19 (10) | |
| | None | 57 (25) | 5 (13) | | 28 (29) | 35 (20) | | 22 (23) | 41 (23) | | 21 (28) | 42 (21) | |
| | Other | 4 (2) | 1 (3) | | 2 (2) | 3 (2) | | 5 (5) | 0 (0) | | 0 (0) | 5 (3) | |
| Method of confirming weight | Self-weighed | 150 (65) | 16 (40) | **<0.001** | 55 (57) | 112 (64) | 0.31 | 61 (63) | 106 (61) | 0.88 | 49 (65) | 118 (60) | 0.47 |
| | Weighed by professional | 74 (32) | 23 (58) | | 38 (39) | 60 (34) | | 34 (35) | 64 (37) | | 24 (32) | 74 (38) | |
| | None | 6 (3) | 1 (3) | | 4 (4) | 3 (2) | | 2 (2) | 5 (3) | | 3 (4) | 4 (2) | |
| If self-weighed, did you have to send proof of weight | Yes | 95 (63) | 12 (75) | 0.35 | 37 (67) | 70 (63) | 0.55 | 44 (72) | 63 (59) | 0.10 | 37 (76) | 70 (59) | **0.047** |
| | No | 55 (37) | 4 (25) | | 18 (33) | 42 (38) | | 17 (28) | 43 (41) | | 12 (25) | 48 (41) | |
| **Experiences of using GLP-1 medication** | | | | | | | | | | | | | |
| GLP1-RA medications are easy to use | Agree | 215 (94) | 36 (90) | 0.37 | 92 (95) | 161 (92) | 0.29 | 90 (93) | 163 (91) | 0.95 | 71 (93) | 182 (93) | 0.92 |
| | Neutral | 7 (3) | 3 (8) | | 4 (4) | 6 (3) | | 4 (4) | 6 (3) | | 3 (4) | 7 (4) | |
| | Disagree | 8 (4) | 1 (3) | | 1 (1) | 8 (5) | | 3 (3) | 6 (3) | | 2 (3) | 7 (4) | |

| | | | | | | | | | | | | | |
|---|---|---|---|---|---|---|---|---|---|---|---|---|---|
| GLP1-RA medications are easy to access | Agree | 175 (76) | 25 (63) | 0.06 | 73 (75) | 128 (73) | 0.93 | 70 (72) | 131 (75) | 0.63 | 61 (80) | 140 (71) | 0.23 |
| | Neutral | 26 (11) | 10 (25) | | 12 (12) | 24 (14) | | 12 (12) | 24 (14) | | 6 (8) | 30 (15) | |
| | Disagree | 29 (13) | 5 (13) | | 12 (12) | 23 (13) | | 15 (16) | 20 (11) | | 9 (12) | 26 (13) | |
| GLP-1 medications improved my overall quality of life | Agree | 130 (57) | 26 (65) | 0.37 | 54 (56) | 103 (59) | 0.85 | 62 (64) | 95 (54) | 0.13 | 36(47) | 121 (62) | 0.09 |
| | Neutral | 49 (21) | 9 (23) | | 21 (22) | 37 (21) | | 21 (22) | 37 (21) | | 19 (25) | 39 (20) | |
| | Disagree | 51 (22) | 5 (13) | | 22 (23) | 35 (20) | | 14 (14) | 43 (25) | | 21 (28) | 36 (18) | |
| I experienced significant side effects while using GLP1-RA medications | Agree | 88 (38) | 14 (35) | 0.73 | 39 (40) | 64 (37) | 0.84 | 32 (33) | 71 (41) | 0.43 | 35 (46) | 68 (35) | 0.11 |
| | Neutral | 45 (20) | 10 (25) | | 19 (20) | 36 (21) | | 20 (21) | 35 (20) | | 10 (13) | 45 (23) | |
| | Disagree | 97 (42) | 97 (42) | | 39 (40) | 75 (43) | | 45 (46) | 69 (39) | | 31 (41) | 83 (42) | |
| Since starting GLP1-RA medication, how has your physical activity level changed? | Increased | 103 (45) | 19 (48) | 0.38 | 45 (46) | 78 (45) | 0.96 | 50 (52) | 73 (42) | 0.22 | 23 (30) | 100 (51) | **<0.01** |
| | Stayed the same | 98 (43) | 19 (48) | | 41 (42) | 77 (44) | | 39 (40) | 79 (45) | | 46 (61) | 72 (37) | |
| | Decreased | 29 (13) | 2 (5) | | 11 (11) | 20 (11) | | 8 (8) | 23 (13) | | 7 (9) | 24 (12) | |
| Since starting GLP1-RAs, how has your mental health changed? | Improved | 94 (41) | 14 (35) | 0.39 | 37 (38) | 73 (42) | 0.85 | 41 (42) | 69 (39) | 0.90 | 30 (40) | 80 (41) | 0.69 |
| | Stayed the same | 107 (47) | 23 (58) | | 48 (50) | 82 (47) | | 45 (46) | 85 (49) | | 35 (46) | 95 (49) | |

| | | | | | | | | | | | | | |
|---|---|---|---|---|---|---|---|---|---|---|---|---|---|
| | Worsened | 29 (13) | 3 (8) | | 12 (12) | 20 (11) | | 11 (11) | 21 (12) | | 11 (15) | 21 (11) | |
| How has the quantity of food you consume been affected? | Less food | 214 (93) | 35 (88) | 0.26 | 89 (92) | 162 (93) | 0.48 | 92 (95) | 159 (91) | 0.38 | 67 (88) | 184 (94) | 0.10 |
| | No change | 15 (7) | 4 (10) | | 8 (8) | 11 (6) | | 5 (5) | 14 (8) | | 9 (12) | 10 (5) | |
| | More food | 1 (<1) | 1 (3) | | 0 (0) | 2 (1) | | 0 (0) | 2 (1) | | 0 (0) | 2 (1) | |
| Do you feel GLP1-RA has improved the quality of your food choices? | Healthier | 151 (66) | 34 (85) | **0.05** | 66 (68) | 120 (69) | 0.91 | 59 (61) | 127 (73) | 0.13 | 45 (59) | 141 (72) | 0.12 |
| | Stayed the same | 70 (30) | 5 (13) | | 28 (29) | 48 (27) | | 33 (34) | 43 (25) | | 28 (37) | 48 (25) | |
| | Less healthy | 9 (4) | 1 (3) | | 3 (3) | 7 (4) | | 5 (5) | 5 (3) | | 3 (4) | 7 (4) | |
| **Healthcare provider support** | | | | | | | | | | | | | |
| My healthcare provider is knowledgeable about GLP-1RA medication | Agree | 150 (65) | 31 (78) | 0.18 | 62 (64) | 120 (69) | 0.52 | 63 (65) | 119 (68) | 0.88 | 41 (54) | 141 (72) | **0.01** |
| | Neutral | 60 (26) | 5 (13) | | 24 (25) | 42 (24) | | 25 (26) | 41 (23) | | 24 (32) | 42 (21) | |
| | Disagree | 20 (9) | 4 (10) | | 11 (11) | 13 (7) | | 9 (9) | 15 (9) | | 11 (15) | 13 (7) | |
| My healthcare provider provides adequate support | Agree | 119 (52) | 28 (70) | 0.08 | 49 (51) | 99 (57) | 0.32 | 53 (55) | 95 (54) | 0.99 | 30 (40) | 118 (60) | **<0.001** |
| | Neutral | 71 (31) | 9 (23) | | 28 (29) | 52 (30) | | 28 (29) | 52 (30) | | 25 (33) | 55 (28) | |
| | Disagree | 40 (17) | 3 (8) | | 20 (21) | 24 (14) | | 16 (17) | 28 (16) | | 21 (28) | 23 (12) | |

| | | | | | | | | | | | | | |
|---|---|---|---|---|---|---|---|---|---|---|---|---|---|
| My healthcare provider provides adequate dietary advice during my treatment | Agree | 106 (46) | 26 (65) | | 40 (41) | 92 (53) | | 41 (42) | 91 (52) | | 26 (34) | 106 (54) | |
| | Neutral | 64 (28) | 8 (20) | 0.08 | 28 (29) | 45 (26) | 0.17 | 29 (30) | 44 (25) | 0.31 | 22 (29) | 51 (26) | **<0.01** |
| | Disagree | 60 (26) | 6 (15) | | 29 (30) | 38 (22) | | 27 (28) | 40 (23) | | 28 (37) | 39 (20) | |
| My healthcare provider provides adequate physical activity guidance | Agree | 80 (35) | 21 (53) | | 31 (32) | 70 (40) | | 35 (36) | 66 (38) | | 17 (22) | 84 (43) | |
| | Neutral | 72 (31) | 10 (25) | 0.10 | 30 (31) | 52 (30) | 0.37 | 31 (32) | 51 (29) | 0.89 | 27 (36) | 55 (28) | **<0.01** |
| | Disagree | 78 (34) | 9 (23) | | 36 (37) | 53 (30) | | 31 (32) | 58 (33) | | 32 (41) | 57 (29) | |
| My healthcare provider provides adequate psychological support | Agree | 55 (24) | 22 (55) | | 21 (23) | 55 (31) | | 23 (24) | 54 (31) | | 12 (16) | 65 (33) | |
| | Neutral | 76 (33) | 6 (15) | **<0.001** | 31 (32) | 51 (29) | 0.30 | 36 (37) | 46 (26) | 0.15 | 23 (30) | 59 (30) | **<0.01** |
| | Disagree | 99 (43) | 12 (30) | | 44 (45) | 69 (30) | | 38 (39) | 75 (43) | | 41 (54) | 71 (37) | |
| My healthcare provider regularly monitors my weight and health | Agree | 115 (50) | 25 (63) | | 45 (46) | 96 (55) | | 52 (54) | 89 (51) | | 31 (41) | 110 (56) | |
| | Neutral | 43 (19) | 9 (23) | 0.11 | 19 (20) | 33 (19) | 0.34 | 17 (18) | 35 (20) | 0.86 | 16 (21) | 36 (18) | 0.06 |
| | Disagree | 72 (31) | 6 (15) | | 33 (34) | 46 (26) | | 28 (29) | 51 (29) | | 29 (38) | 50 (26) | |
| I feel comfortable discussing my concerns with my healthcare provider | Agree | 149 (65) | 31 (78) | 0.25 | 62 (64) | 119 (68) | 0.67 | 62 (64) | 119 (68) | 0.79 | 42 (55) | 139 (71) | **0.04** |
| | Neutral | 55 (24) | 7 (18) | | 23 (24) | 40 (23) | | 24 (25) | 39 (22) | | 22 (29) | 41 (21) | |

| | | | | | | | | | | | | | |
|---|---|---|---|---|---|---|---|---|---|---|---|---|---|
| | Disagree | 26 (11) | 2 (5) | | 12 (12) | 16 (9) | | 11 (11) | 17 (10) | | 12 (16) | 16 (8) | |
| **Barriers to adherence** | | | | | | | | | | | | | |
| **Cost of medication** | Extremely challenging | 77 (34) | 11 (28) | 0.15 | 36 (37) | 53 (30) | 0.33 | 41 (42) | 48 (27) | **0.04** | 30 (40) | 59 (30) | 0.08 |
| | Moderately challenging | 74 (32) | 20 (50) | | 27 (28) | 68 (39) | | 26 (27) | 69 (39) | | 19 (25) | 76 (39) | |
| | Slightly challenging | 34 (15) | 5 (13) | | 15 (16) | 24 (14) | | 11 (11) | 28 (16) | | 9 (12) | 30 (15) | |
| | Not challenging | 45 (20) | 4 (10) | | 19 (20) | 30 (17) | | 19 (20 | 30 (17) | | 19 (24) | 31 (16) | |
| **Side effects** | Extremely challenging | 52 (23) | 6 (15) | 0.25 | 19 (20) | 40 (23) | 0.12 | 15 (16) | 44 (25) | 0.08 | 21 (28) | 38 (19) | 0.27 |
| | Moderately challenging | 40 (17) | 11 (28) | | 20 (21) | 31 (18) | | 22 (23) | 29 (17) | | 11 (15) | 40 (20) | |
| | Slightly challenging | 66 (29) | 14 (35) | | 22 (23) | 59 (34) | | 25 (26) | 56 (32) | | 19 (25) | 61 (32) | |
| | Not challenging | 72 (31) | 9 (23) | | 36 (37) | 45 (26) | | 35 (36) | 46 (26) | | 25 (33) | 56 (29) | |
| **Access to medication** | Extremely challenging | 19 (8) | 3 (8) | 0.06 | 8 (8) | 22 (8) | 0.20 | 10 (10) | 12 (7) | 0.31 | 8 (11) | 14 (7) | 0.09 |
| | Moderately challenging | 29 (13) | 11 (28) | | 13 (13) | 28 (16) | | 11 (11) | 30 (17) | | 8 (11) | 33 (17) | |
| | Slightly challenging | 48 (21) | 10 (25) | | 15 (16) | 44 (25) | | 18 (19) | 41 (23) | | 11 (15) | 48 (25) | |
| | Not challenging | 134 (58) | 16 (40) | | 61 (63) | 89 (51) | | 58 (60) | 92 (53) | | 49 (65) | 101 (52) | |
| **Social stigma** | Extremely challenging | 10 (4) | 2 (5) | 0.28 | 4 (4) | 8 (5) | 0.50 | 5 (5) | 7 (4) | 0.05 | 3 (4) | 9 (5) | 0.56 |
| | Moderately challenging | 22 (10) | 5 (13) | | 7 (7) | 21 (12) | | 4 (4) | 24 (14) | | 7 (9) | 21 (11) | |

| | | | | | | | | | | | | | |
|---|---|---|---|---|---|---|---|---|---|---|---|---|---|
| | Slightly challenging | 47 (20) | 13 (33) | | 25 (26) | 35 (20) | | 19 (20) | 41 (23) | | 13 (17) | 47 (24) | |
| | Not challenging | 151 (66) | 20 (50) | | 61 (63) | 111 (63) | | 69 (71) | 103 (59) | | 53 (70) | 119 (61) | |
| **Negative effects on social life** | Extremely challenging | 7 (3) | 3 (8) | 0.22 | 4 (4) | 6 (3) | 0.66 | 3 (3) | 7 (4) | 0.88 | 4 (5) | 6 (3) | 0.83 |
| | Moderately challenging | 28 (12) | 8 (20) | | 10 (10) | 26 (15) | | 11 (11) | 25 (14) | | 9 (12) | 27 (14) | |
| | Slightly challenging | 61 (27) | 11 (28) | | 29 (30) | 44 (25) | | 27 (28) | 46 (26) | | 20 (26) | 53 (27) | |
| | Not challenging | 134 (58) | 18 (45) | | 54 (56) | 99 (57) | | 56 (58) | 97 (55) | | 43 (57) | 110 (56) | |
| **Motivators to adherence** | | | | | | | | | | | | | |
| **Improved health outcomes** | Extremely important | 110 (48) | 25 (63) | 0.18 | 45 (46) | 91 (52) | 0.84 | 50 (52) | 86 (49) | 0.88 | 35 (46) | 101 (52) | 0.61 |
| | Moderately important | 82 (36) | 13 (33) | | 36 (37) | 60 (34) | | 32 (33) | 64 (37) | | 28 (37) | 68 (35) | |
| | Slightly important | 29 (13) | 1 (3) | | 12 (12) | 18 (10) | | 12 (12) | 18 (10) | | 11 (15) | 19 (10) | |
| | Not important | 9 (4) | 1 (3) | | 4 (4) | 6 (3) | | 3 (3) | 7 (4) | | 2 (3) | 8 (4) | |
| **Weight loss** | Extremely important | 186 (81) | 29 (73) | 0.18 | 81 (84) | 136 (78) | 0.20 | 78 (80) | 139 (79) | 0.73 | 64 (84) | 153 (78) | **0.047** |
| | Moderately important | 35 (15) | 11 (28) | | 12 (12) | 34 (19) | | 16 (17) | 30 (17) | | 8 (11) | 38 (19) | |
| | Slightly important | 7 (3) | 0 (0) | | 4 (4) | 3 (2) | | 3 (3) | 4 (2) | | 2 (3) | 5 (3) | |
| | Not important | 2 (1) | 0 (0) | | 0 (0) | 2 (1) | | 0 (0) | 2 (1) | | 2 (3) | 0 (0) | |
| **Improved body image** | Extremely important | 117 (51) | 25 (63) | 0.45 | 53 (55) | 91 (52) | 0.94 | 46 (47) | 98 (56) | 0.32 | 44 (58) | 100 (51) | 0.08 |

| | | | | | | | | | | | | | |
|---|---|---|---|---|---|---|---|---|---|---|---|---|---|
| | Moderately important | 76 (33) | 12 (30) | | 31 (32) | 57 (33) | | 32 (33) | 56 (32) | | 21 (28) | 67 (34) | |
| | Slightly important | 24 (10) | 2 (5) | | 9 (9) | 17 (10) | | 13 (13) | 13 (7) | | 4 (5) | 22 (11) | |
| | Not important | 13 (6) | 1 (3) | | 4 (4) | 10 (6) | | 6 (6) | 8 (5) | | 7 (9) | 7 (4) | |
| **Increased self confidence** | Extremely important | 106 (46) | 18 (45) | 0.76 | 49 (51) | 77 (44) | 0.30 | 43 (44) | 83 (47) | 0.24 | 37 (49) | 89 (45) | 0.83 |
| | Moderately important | 83 (36) | 17 (43) | | 36 (37) | 64 (37) | | 35 (36) | 65 (37) | | 26 (34) | 74 (38) | |
| | Slightly important | 28 (12) | 4 (10) | | 10 (10) | 22 (13) | | 16 (17) | 16 (9) | | 8 (11) | 24 (12) | |
| | Not important | 13 (6) | 1 (3) | | 2 (2) | 12 (7) | | 3 (3) | 11 (6) | | 5 (7) | 9 (5) | |
| **Improved quality of life** | Extremely important | 111 (48) | 21 (53) | 0.27 | 46 (47) | 88 (50) | 0.29 | 49 (51) | 85 (49) | 0.97 | 36 (47) | 98 (50) | 0.83 |
| | Moderately important | 74 (32) | 16 (40) | | 36 (37) | 54 (31) | | 31 (32) | 59 (34) | | 28 (37) | 62 (32) | |
| | Slightly important | 35 (15) | 3 (8) | | 14 (14) | 24 (14) | | 14 (14) | 24 (14) | | 10 (13) | 28 (14) | |
| | Not important | 10 (4) | 0 (0) | | 1 (1) | 9 (5) | | 3 (3) | 7 (4) | | 2 (3) | 8 (4) | |
| **Improved social life** | Extremely important | 34 (15) | 14 (35) | **0.01** | 17 (18) | 31 (18) | 0.42 | 13 (13) | 35 (20) | 0.40 | 10 (13) | 38 (19) | 0.65 |
| | Moderately important | 44 (19) | 10 (25) | | 19 (20) | 36 (21) | | 20 (21) | 35 (20) | | 17 (22) | 38 (19) | |
| | Slightly important | 61 (27) | 7 (18) | | 30 (31) | 39 (22) | | 23 (24) | 46 (26) | | 19 (25) | 50 (26) | |
| | Not important | 91 (40) | 9 (23) | | 31 (32) | 69 (39) | | 41 (42) | 59 (34) | | 30 (40) | 70 (36) | |
| **Improved pain management** | Extremely important | 34 (15) | 8 (20) | 0.55 | 17 (18) | 26 (15) | 0.12 | 13 (13) | 30 (17) | 0.11 | 15 (20) | 28 (14) | 0.33 |

| | | | | | | | | | | | | | |
|---|---|---|---|---|---|---|---|---|---|---|---|---|---|
| | Moderately important | 42 (18) | 9 (23) | | 24 (25) | 28 (16) | | 26 (27) | 26 (15) | | 18 (24) | 34 (17) | |
| | Slightly important | 40 (17) | 8 (20) | | 19 (20) | 29 (17) | | 15 (16) | 33 (19) | | 11 (15) | 37 (19) | |
| | Not important | 114 (50) | 15 (38) | | 37 (38) | 92 (53) | | 43 (44) | 86 (49) | | 32 (42) | 97 (50) | |
| **Improved physical activity** | Extremely important | 49 (21) | 11 (28) | 0.21 | 18 (19) | 43 (25) | 0.28 | 17 (18) | 44 (25) | 0.34 | 12 (16) | 49 (25) | 0.20 |
| | Moderately important | 83 (36) | 19 (48) | | 43 (44) | 60 (34) | | 43 (44) | 60 (34) | | 27 (36) | 76 (39) | |
| | Slightly important | 53 (23) | 6 (15) | | 22 (23) | 37 (21) | | 20 (21) | 39 (22) | | 19 (25) | 40 (20) | |
| | Not important | 45 (20) | 4 (10) | | 14 (14) | 35 (20) | | 17 (18) | 32 (18) | | 18 (24) | 31 (16) | |
| **Improved diet quality** | Extremely important | 65 (28) | 17 (43) | 0.06 | 32 (33) | 51 (29) | 0.78 | 28 (29) | 55 (31) | 0.86 | 19 (25) | 64 (33) | 0.43 |
| | Moderately important | 78 (34) | 16 (40) | | 34 (35) | 60 (34) | | 32 (33) | 62 (35) | | 30 (40) | 64 (33) | |
| | Slightly important | 54 (24) | 6 (15) | | 21 (22) | 39 (22) | | 24 (25) | 36 (21) | | 15 (20) | 45 (23) | |
| | Not important | 33 (14) | 1 (3) | | 10 (10) | 25 (14) | | 13 (13) | 22 (13) | | 12 (16) | 23 (12) | |
| **Reduction in other medication** | Extremely important | 14 (6) | 6 (15) | 0.10 | 8 (8) | 12 (7) | 0.48 | 8 (8) | 12 (7) | 0.96 | 7 (9) | 13 (7) | 0.69 |
| | Moderately important | 34 (15) | 9 (23) | | 19 (20) | 24 (14) | | 16 (17) | 27 (15) | | 14 (18) | 29 (15) | |
| | Slightly important | 32 (14) | 5 (13) | | 15 (16) | 24 (14) | | 14 (14) | 25 (14) | | 9 (12) | 30 (15) | |
| | Not important | 150 (65) | 20 (50) | | 55 (57) | 115 (66) | | 59 (61) | 111 (63) | | 46 (61) | 124 (63) | |
| **Discontinuation and post-treatment impact** | | | | | | | | | | | | | |

| | | | | | | | | | | | | | |
|---|---|---|---|---|---|---|---|---|---|---|---|---|---|
| **Reason for stopping GLP-1 medication** | Cost of treatment | 72 (31) | 10 (25) | 0.29 | 32 (33) | 51 (29) | 0.80 | 33 (34) | 50 (29) | 0.27 | 23 (30) | 60 (31) | 0.07 |
| | Planned end of treatment | 52 (23) | 52 (23) | | 23 (24) | 44 (25) | | 21 (22) | 33 (34) | | 10 (13) | 57 (29) | |
| | Side effects | 61 (27) | 61 (27) | | 25 (26) | 44 (25) | | 23 (24) | 46 (26) | | 25 (33) | 44 (22) | |
| | Lack of motivation/benefit | 9 (4) | 3 (8) | | 2 (2) | 10 (6) | | 3 (3) | 9 (5) | | 5 (7) | 7 (4) | |
| | Accessibility issues | 6 (3) | 1 (3) | | 3 (3) | 4 (2) | | 5 (5) | 2 (1) | | 3 (4) | 4 (2) | |
| | Stigma | 2 (1) | 1 (3) | | 2 (2) | 1 (1) | | 0 (0) | 3 (2) | | 2 (3) | 1 (1) | |
| | Changes to social life | 2 (1) | 1 (3) | | 1 (1) | 2 (1) | | 2 (2) | 1 (1) | | 0 (0) | 3 (2) | |
| | Other | 26 (11) | 2 (5) | | 9 (9) | 19 (11) | | 10 (10) | 18 (10) | | 8 (11) | 20 (10) | |
| **Barriers to restarting GLP-1 medication** | Cost of treatment | 78 (34) | 11 (28) | 0.29 | 37 (38) | 53 (30) | 0.84 | 40 (41) | 50 (29) | 0.33 | 28 (37) | 62 (32) | 0.11 |
| | Planned end of treatment | 55 (24) | 14 (35) | | 23 (24) | 46 (26) | | 20 (21) | 49 (28) | | 11 (15) | 58 (30) | |

| | | | | | | | | | | | | | |
|---|---|---|---|---|---|---|---|---|---|---|---|---|---|
| | Side effects | 53 (23) | 6 (15) | | 19 (20) | 40 (23) | | 18 (19) | 41 (23) | | 19 (25) | 40 (20) | |
| | Lack of motivation/benefit | 20 (9) | 6 (15) | | 10 (10) | 16 (9) | | 7 (7) | 19 (11) | | 9 (12) | 17 (9) | |
| | Accessibility issues | 10 (4) | 1 (3) | | 3 (3) | 8 (5) | | 4 (4) | 7 (4) | | 2 (3) | 9 (5) | |
| | Stigma | 0 (0) | 0 (0) | | 0 (0) | 0 (0) | | 0 (0) | 0 (0) | | 0 (0) | 0 (0) | |
| | Changes to social life | 1 (<1) | 1 (3) | | 1 (1) | 1 (1) | | 1 (1) | 1 (1) | | 0 (0) | 2 (1) | |
| | Other | 13 (6) | 1 (3) | | 4 (4) | 11 (6) | | 7 (7) | 8 (5) | | 7 (9) | 8 (4) | |
| **What happened to your weight after you stopped taking GLP-1 medication?** | Gained weight | 108 (47) | 13 (33) | 0.20 | 41 (42) | 82 (47) | 0.28 | 41 (42) | 82 (47) | 0.51 | 40 (53) | 83 (42) | 0.31 |
| | Maintained | 89 (39) | 21 (53) | | 45 (46) | 65 (37) | | 39 (40) | 71 (41) | | 27 (36) | 83 (42) | |
| | Lost weight | 33 (14) | 6 (15) | | 11 (11) | 28 (16) | | 17 (18) | 22 (13) | | 9 (12) | 30 (15) | |
| **What happened to your other health conditions after** | Improvement | 29 (22) | 10 (40) | 0.12 | 10 (17) | 29 (29) | 0.25 | 12 (22) | 27 (26) | 0.54 | 11 (22) | 28 (25) | 0.69 |

| | | | | | | | | | | | | | |
|---|---|---|---|---|---|---|---|---|---|---|---|---|---|
| **you stopped taking GLP-1 medication?** | No change | 92 (69) | 14 (56) | | 43 (73) | 63 (62) | | 36 (66) | 70 (67) | | 32 (65) | 74 (67) | |
| | Worsening | 13 (10) | 1 (4) | | 6 (10) | 9 (9) | | 7 (13) | 8 (8) | | 6 (12) | 9 (8) | |
| **What happened to your physical activity after you stopped taking GLP-1 medication?** | Increased | 17 (34) | 14 (35) | 0.37 | 25 (26) | 66 (38) | 0.09 | 30 (31) | 61 (35) | 0.63 | 14 (18) | 77 (39) | **<0.01** |
| | No change | 132 (57) | 25 (63) | | 61 (63) | 97 (55) | | 57 (59) | 101 (58) | | 52 (68) | 106 (54) | |
| | Decreased | 21 (9) | 1 (3) | | 11 (11) | 12 (7) | | 10 (10) | 13 (7) | | 10 (13) | 13 (7) | |
| **What happened to your mental health after you stopped taking GLP-1 medication?** | Improvement | 47 (20) | 12 (30) | 0.37 | 17 (18) | 42 (24) | 0.46 | 21 (22) | 38 (22) | 0.95 | 12 (16) | 47 (24) | 0.08 |
| | No change | 144 (63) | 23 (58) | | 63 (65) | 105 (60) | | 59 (61) | 109 (62) | | 46 (61) | 122 (62) | |
| | Worsening | 39 (17) | 5 (13) | | 17 (18) | 28 (16) | | 17 (18) | 28 (16) | | 18 (24) | 27 (14) | |
| **What happened to the quantity of food you consumed when you stopped taking GLP-1 medication?** | Increased | 39 (17) | 17 (43) | <0.001 | 16 (17) | 40 (23) | 0.35 | 20 (21) | 36 (21) | 0.54 | 9 (12) | 47 (24) | 0.08 |
| | No change | 54 (24) | 9 (23) | | 26 (27) | 37 (21) | | 26 (27) | 37 (21) | | 19 (25) | 44 (22) | |

| | | | | | | | | | | | | | |
|---|---|---|---|---|---|---|---|---|---|---|---|---|---|
| | Decreased | 137 (60) | 14 (35) | | 55 (57) | 98 (56) | | 51 (53) | 102 (58) | | 48 (63) | 105 (54) | |
| **What happened to the quality of food you consumed when you stopped taking GLP-1 medication?** | Increased | 76 (33) | 19 (48) | 0.20 | 27 (28) | 68 (39) | 0.19 | 27 (28) | 68 (39) | 0.19 | 20 (26) | 75 (38) | **0.02** |
| | No change | 123 (54) | 16 (40) | | 55 (57) | 85 (49) | | 55 (57) | 85 (49) | | 39 (51) | 101 (52) | |
| | Decreased | 31 (14) | 5 (13) | | 15 (16) | 22 (13) | | 15 (16) | 22 (13) | | 17 (22) | 20 (10) | |

**Supplementary material 9 – Full reporting of subgroup differences by demographics**

Subgroup analyses by age, gender, BMI (at time of survey and treatment), ethnicity, social status, education and exercise are summarised below.

**Age**

Among current users, younger participants reported longer treatment (1–2 years: 15% vs. 8%, $p=0.02$) and preferred private online access (80% vs. 73%), whereas older users more often accessed NHS face-to-face services (19% vs. 12% ($p=0.04$). Younger users emphasised the importance of body image (76% vs. 48%), self-confidence (65% vs. 43%), and social life (26% vs. 9%) for adherence motivators, while older users emphasized improved health (74% vs. 61%).

Among former users, younger participants preferred private online access (64% vs. 51%, $p<0.001$) and self-weighed for treatment eligibility (69% vs. 52%, $p=0.02$). Comparisons between age groups in former users for other responses were generally similar ($p>0.05$).

**Gender**

Female current users preferred to access medication privately online (83% vs 57%, $p<0.001$) and typically self-weighed for eligibility (83% vs 55%, $p<0.001$), while males more commonly used NHS face-to-face services (31% vs. 10%) and had weight confirmed by a professional (43% vs. 15%). Females reported easier access to medication (73% vs. 63%, $p=0.03$). Females prioritised motivators to adherence such as weight loss (90% vs 78%, $p<0.01$), self-confidence (59% vs. 40%, $p=0.03$), pain management (27% vs 12%, $p<0.01$) and physical activity (28% vs. 15%, $p=0.048$).

Former female users again preferred private online access (68% vs 36%, $p<0.001$) and typically self-weighed for eligibility (72% vs 39%) (both $p<0.001$), while males used NHS face-to-face services more (41% vs 15%) and had weight confirmed by a professional (57% vs 26%). Females reported more side effects (44% vs 24%, $p<0.001$) and reduced food intake (95% vs 87%, $p=0.03$), while males more frequently reported improved quality of life (70% vs 52%, $p=0.02$) and better healthcare experiences. Females cited cost (41% vs 13%, $p<0.001$) and side effects (26% vs 12%, $p=0.051$) as major adherence barriers, and valued weight loss (86% vs 66%), body image (61% vs 35%), self-confidence (54% vs 31%) and quality of life (55% vs 37%) as motivators (all $p\leq0.04$). Males more often prioritised medication reduction (12% vs

5%, $p$=0.02). Females tended to discontinue to a greater extent than males as a result of cost (38% vs 14%) and side effects (31% vs 12%), while more males stopped for planned completion (50% vs 13%) ($p$<0.001). Post-discontinuation, females reported greater prevalence of weight gain (52% vs 29%, $p$<0.001), whereas males reported greater improvements in other health conditions (36% vs 18%), physical activity (46% vs 28%), mental health (30% vs 18%) and increased food quantity (38% vs 13%) and quality (46% vs 30%) (all $p \leq 0.03$).

**BMI at time of survey**

In current users, those with a lower BMI at the time of survey reported greater improvements in quality of life (94% vs. 81%, $p$<0.001), physical activity (65% vs. 60%, $p$=0.02) and mental health (63% vs. 55%, $p$=0.046). They also reported better psychological support from healthcare providers than those with a higher BMI (33% vs. 23%, $p$=0.01). Those with a higher BMI more frequently identified improved health outcomes (76% vs. 57%, $p$<0.01) and weight loss (94% vs. 74%, $p$<0.001) as extremely important motivators.

In former users, those with a lower BMI more frequently reported improvements in quality of life (67% vs. 43%, $p$<0.001), physical activity (52% vs. 34%, $p$=0.01), and diet quality (75% vs. 57%, $p$<0.001), and rated healthcare support more positively, including adequate support (62% vs. 42%, $p$<0.001), dietary advice (56% vs. 37%, $p$<0.001), physical activity guidance (43% vs. 28%, $p$<0.001), psychological support (35% vs. 18%, $p$<0.001), and regular monitoring (59% vs. 41%, $p$=0.02). Barriers also differed, as those with a higher BMI more often rated cost (40% vs. 29%, $p$=0.04) and side effects (32% vs. 16%, $p$=0.01) as extremely challenging, while also more often reporting access as not challenging (63% vs. 50%, $p$=0.03). In terms of motivators, pain management was more often rated extremely important by those with a higher BMI (23% vs. 12%, $p$=0.03). Discontinuation reasons differed, with participants of a lower BMI mainly stopping due to planned treatment completion (33% vs. 1%), whereas participants with a higher BMI more frequently stopping due to side effects (31% vs. 12%) or cost (38% vs. 14%) ($p$<0.001). After discontinuation, participants with a lower BMI more often increased food quantity (28% vs. 8%, $p$<0.001) and quality (42% vs. 24%, $p$=0.01).

**BMI at start of treatment**

Current users with a higher BMI at the start of treatment more frequently disagreed that dietary advice (28% vs. 19%, $p$=0.03) and psychological support (50% vs. 34%, $p$=0.01) were adequate and identified cost as an extremely challenging barrier to adherence (40% vs. 25%, $p=0.02$). In terms of motivators, participants with a higher BMI more often rated improved health outcomes (78% vs. 61%, $p=0.01$), weight loss (93% vs. 84%, $p<0.001$), pain management (28% vs. 21%, $p<0.001$), and diet quality (36% vs. 31%, $p=0.03$) as extremely important.

In former users, those with a lower BMI at the start of treatment were more likely to access medication privately online (60% vs. 52%, $p=0.02$) and to confirm weight through self-weighing (66% vs. 49%, $p=0.03$). By contrast, participants with a higher BMI more frequently reported access as extremely challenging (17% vs. 5%, $p<0.001$). Pain management was also more often identified as an extremely important motivator among those with higher BMI (22% vs. 14%, $p<0.001$).

**Ethnicity**

In current users, white participants were more likely to use Tirzepatide (81% vs. 73%, $p=0.03$), access privately online (79% vs. 61%, $p=0.054$), and self-weigh for treatment monitoring (80% vs. 51%, $p<0.001$), whereas participants from other ethnic backgrounds more often had weight confirmed by a professional (44% vs. 19%). White participants also reported greater ease of use (97% vs. 90%, $p<0.01$), comfort discussing concerns with healthcare provider (85% vs. 70%, $p=0.03$) and reduced food intake (97% vs. 90%, $p<0.001$). Participants from other ethnicities placed more importance on improvements in social life (32% vs. 15%, $p<0.001$) and reduction of other medications (27% vs. 12%, $p=0.02$).

In former users, white participants more often used private online access (62% vs. 38%, $p<0.001$) and self-weighed for eligibility (65% vs. 40%, $p<0.001$), while participants from other backgrounds more frequently had weight confirmed by a professional (58% vs. 32%). Participants from other ethnicities also reported greater psychological support (55% vs. 24%, $p<0.001$), improved quality of food intake (85% vs. 66%, $p=0.05$), and placed higher importance on social life improvements as a motivator to adherence (35% vs. 15%, $p=0.01$). Post-treatment, participants from other ethnicities were more likely to increase the quantity of food consumed (43% vs. 17%, $p<0.001$).

**Social status**

Among current users, those with a self-reported higher social status more often reported that GLP-1 medications were easy to use (98% vs. 92%, $p$=0.02) and easy to access (74% vs. 65%, $p$<0.001).

Among former users, participants with a lower social status were more likely to access medication via NHS face-to-face services (30% vs. 19%, $p$=0.01), while participants with higher social status were more likely to have their treatment managed privately (47% vs. 37%, $p$=0.02). Barriers also differed by social status, as participants with a lower social status reported cost as an extremely challenging factor more often (42% vs. 27%, $p$=0.04), whereas participants with a higher social status were more likely to rate social stigma as moderately challenging (14% vs. 4%, $p$=0.05).

**Exercise**

Current users who did not engage in regular exercise were more likely to be required to provide proof of weight for GLP-1 treatment (76% vs. 59%, $p$=0.047). GLP-1 use was associated with greater increases in physical activity among those who already exercised (51% vs. 30%, $p$=0.01). Participants who engage in exercise also reported higher agreement that their healthcare provider was knowledgeable (72% vs. 54%, $p$=0.01), provided adequate overall support (60% vs. 40%, $p$<0.001), dietary advice (54% vs. 34%, $p$=0.01), physical activity guidance (43% vs. 22%, $p$<0.01), and psychological support (33% vs. 16%, $p$<0.01), and felt more comfortable discussing concerns with healthcare providers(71% vs. 55%, $p$=0.04). Regarding motivators, participants who did not exercise were more likely to rate weight loss as extremely important (84% vs. 78%, $p$=0.047).

Former users who engaged in regular exercise were more likely to access medication privately online (79% vs. 71%, $p$=0.02) and those who did not engage with exercise more frequently had their treatment managed by their GP (24% vs. 12%, $p$=0.02) and were more likely to have to provide proof of weight (76% vs. 59%, $p$=0.047). Those that did engage with exercise were more likely to report improvements in quality of life (92% vs. 76%, $p$<0.001), physical activity (72% vs. 41%, $p$<0.001), and mental health (63% vs. 47%, $p$=0.02). GLP-1 use increased physical activity more in those who exercised (51% vs. 30%, $p$=0.01), and improved physical activity was also more often rated as an extremely important motivator in this group (30% vs. 15%, $p$<0.001). Those who did not exercise reported that social stigma was not challenging (58% vs. 43%, $p$=0.02). Post-treatment, those who exercised were more likely to report increased physical activity (39% vs. 18%, $p<0.01$) and improvements in diet quality (38% vs. 26%, $p=0.02$).
